# An agentic AI system enhances clinical detection of immunotherapy toxicities: a multi-phase validation study

**DOI:** 10.64898/2026.02.26.26347179

**Authors:** Jack Gallifant, Shan Chen, Kee-Young Shin, Katherine C. Kellogg, Patrick F. Doyle, Joyce Guo, Bingyang Ye, Andrew Warrington, Bingxue K Zhai, Matthew J. Hadfield, Alexander Gusev, Biagio Ricciuti, David C. Christiani, Hugo JWL Aerts, Benjamin H. Kann, Raymond H. Mak, Tanna L. Nelson, Paul Nguyen, Jonathan D. Schoenfeld, Umit Topaloglu, Paul Catalano, Harry Hochheiser, Jeremy L. Warner, Elad Sharon, David E. Kozono, Guergana K. Savova, Danielle S. Bitterman

## Abstract

Immune-related adverse events (irAEs) affect up to 40% of patients receiving immune checkpoint inhibitors, yet their identification depends on laborious and inconsistent manual chart review. Here we developed and evaluated an agentic large language model system to extract the presence, temporality, severity grade, attribution, and certainty of six irAE types from clinical notes. Retrospectively (263 notes), the system achieved macro-averaged F1 of 0.92 for detection and 0.66 for multi-class severity grading; self-consistency improved F1 by 0.14. The best-performing configuration cost approximately $0.02 per note. In prospective silent deployment over three months (884 notes), detection F1 was 0.72–0.79. In a randomized crossover study of clinical trial staff (17 participants, 316 observations), agentic assistance reduced annotation time by 40% (P < 0.001), increased complete-match accuracy (OR 1.45; 95% CI 1.01–2.09; P = 0.045), and improved inter-annotator agreement (Krippendorff’s α from 0.22–0.51 to 0.82–0.85). These results demonstrate that agentic AI coupled with human verification could enhance efficiency, performance, and consistency for irAE assessment.

## Introduction

Treatment-emergent adverse events (TEAEs) are under-recognized in clinical trials and existing databases.^1–8^ Accurate detection and reporting of TEAEs is foundational to cancer care, pharmacovigilance^9^, and survivorship planning. In routine practice, however, many clinically important adverse events are documented primarily in unstructured clinical narratives; structured electronic health record (EHR) fields incompletely capture their occurrence, necessitating laborious, resource-intensive, and error-prone manual chart review.^10,11^ This manual bottleneck currently limits the capacity and quality of adverse event curation and monitoring for clinical trials, real-world evidence, and clinical decision-support at the point-of-care.

Against this backdrop, immune checkpoint inhibitors (ICI) have transformed cancer treatment, yet their success comes with a critical challenge: up to 40% of patients develop immune-related adverse events (irAEs) that can affect any organ system.^12–16^ While timely detection of irAEs saves lives, evidence of these toxicities are often buried within clinical notes, escaping standard surveillance methods.^15,17^ Furthermore, addressing the challenge of irAE surveillance and management will become increasingly urgent as the number of cancer survivors grows to an estimated 26 million survivors anticipated by 2040 in the US alone.^18^ Furthermore, the clinical burden is compounded by substantial economic costs, with irAE-related hospitalizations and treatment adding thousands of dollars per patient and requiring resource-intensive manual chart review for detection^19^.

Despite their clinical significance, systematically identifying cancer TEAEs presents a considerable challenge for both clinical care and pharmacovigilance.^20^ Directly analyzable structured data fields within EHRs do not reliably capture irAEs, especially those that do not result in hospitalization but still have important impacts on patients’ overall morbidity and quality of life.^11^ Even of those that result in hospitalization, ICD codes have a recall/sensitivity of only approximately 68%.^21^ Because of this, manual chart curation is required for irAE reporting, but this suffers from the aforementioned manual curation bottleneck and introduces significant quality challenges due to ambiguous definitions and thresholds for irAE occurrence, severity, and attribution.^22,23^

To address these challenges, natural language processing (NLP), which refers to computational analysis of unstructured text and is a central pillar in modern Artificial Intelligence (AI),^24^ has been explored to extract irAEs from EHR documents automatically. Initial efforts using traditional machine learning or small neural language models demonstrated promise, but require large annotated datasets for task-specific training and tuning.^25,26,27^ More recently, large language models (LLMs) have been shown to extract clinical information without extensive fine-tuning in a zero-shot or few-shot scenario through sophisticated prompting. Preliminary studies applying LLMs to irAE identification have demonstrated promise;^21,28,29^ however, they have focused on binary classification (i.e., irAE present/absent) over an entire inpatient admission record or patient chart. Because these approaches operate at the chart or encounter level rather than on individual clinical notes, they are poorly suited for real-time detection and ongoing monitoring. Furthermore, they have not addressed the extraction of irAE clinical attributes necessary to inform clinical decision-making, translational research, and regulatory reporting for clinical trials. These critical attributes include the temporal status (distinguishing between current, active events and historical ones), severity grading (aligned with standardized scales), attribution (the clinician’s assessment of the likelihood that the event is ICI-related), and the certainty of that attribution. Extracting these details accurately from narrative text remains an unmet need to understand and address irAEs and adverse events more broadly.

Concurrently, advanced autonomous prompting strategies and architectures, commonly referred to as “agentic” systems, aim to further enhance LLM capabilities.^30^ These systems decompose complex reasoning tasks into smaller, manageable sub-problems, often addressed by multiple specialized LLM-based or utilizing tools (agents), and with mechanisms to evaluate and synthesize the final output.^30^ It remains an open question whether such agentic architectures can be effectively leveraged for the complex, multi-faceted task of extracting detailed irAE attributes from clinical notes. Even if technically feasible, the impact of integrating such sophisticated AI assistance into ’ real-world clinical and research workflows, including its effect on efficiency, performance, cognitive load, and overall experience, remains unevaluated. Understanding these human-computer interaction (HCI) factors is necessary to translate technological potential into practical, usable, and valuable healthcare applications.

In this study, we evaluated the impact of real-time agentic LLM assistance on human curation of detailed irAEs from clinical notes. We developed an agentic pipeline that extracts not only the presence of six irAEs spanning common and lethal events (myocarditis, dermatitis, thyroiditis, hepatitis, colitis, and pneumonitis),^31^ but also their temporal status, severity grade, ICI attribution, and attribution certainty at the encounter level. We integrated this system into an annotation interface and conducted human-computer interaction studies to determine whether AI assistance improves reviewer efficiency, accuracy, and consistency compared to unaided manual review. We also approximated the costs of computation. We demonstrated the potential of agentic AI to improve both the quality and efficiency of irAE curation, which could enhance our understanding and optimization of the benefits and risks of cancer treatment via clinical trials and real-world evidence generation.

## Results

Figure 3 summarizes the main results from each phase of the study.

### Phase 1: Note-Level irAE System Development and Evaluation

To identify the optimal configuration for note-level irAE extraction (Figure 1), we systematically benchmarked combinations of proprietary and open-source LLMs across agentic and non-agentic architectures on 263 expert-annotated clinical notes. Two design features are central to interpreting these results. First, each irAE was classified by event temporality, that is, whether the clinical note described it as a currently active event or as a historical (past) event, and performance was evaluated for each timeframe separately. Second, to reduce the variability inherent in single LLM outputs, we implemented a self-consistency mechanism in which specific stages of the agentic process were run three times independently, and a judge agent resolved disagreements before proceeding to the next stage; a single-inference variant that omitted this step served as the primary comparator for ablation analyses (Figure 2). Among all model-architecture combinations evaluated, GPT-4.1-mini with the default agentic architecture achieved the strongest overall performance (Appendix Table 1). This configuration incurred an inference cost of $0.021 per note; costs for all model–architecture combinations are reported in Appendix Table 1. For note-level irAE detection, the system attained a temporality-agnostic macro-averaged F1 of 0.93 across the six conditions (Table 1). Performance was consistent when stratified by event temporality: F1 = 0.92 for active (current) events and 0.91 for resolved (past) events. Current temporality detection performance was highest for Hepatitis and Pneumonitis (F1 = 0.96), and lowest for myocarditis (F1 = 0.86). Condition-specific binary classification performance (precision, recall, F1) is presented in Appendix Figure 1.

**Figure 1.** Multi-dimensional validation framework for an agentic LLM-based irAE detection system. 1a, Retrospective development workflow progressing from dataset curation (289 expert-annotated notes) through agent development with prompt optimization on pneumonitis subset (n=26) to validation on held-out test set (n=263), achieving F1 scores of 0.92 (detection), 0.66 (CTCAE grading), and 0.95 (attribution). The agentic architecture employs six specialized agents with self-consistency mechanisms for robust extraction. 1b, Three parallel real-world validation streams evaluating complementary aspects of system performance: prospective pilot demonstrates real-time monitoring capability with 98% agreement (n=1000), controlled HCI study quantifies efficiency gains using crossover design (n=20 notes, 20 CRCs), and field study confirms operational effectiveness with 73% time reduction in chart curation. Total validation encompassed real-world cases across diverse clinical contexts. BWH, Brigham and Women’s Hospital; DFCI, Dana-Farber Cancer Institute; CTCAE, Common Terminology Criteria for Adverse Events; CRC, clinical research coordinator; HCI, human-computer interaction; ICI, immune checkpoint inhibitor; irAE, immune-related adverse event.

**Figure 2.** Multi-stage agentic architecture for irAE detection, grading, attribution, and certainty assessment. Unstructured clinical notes from the electronic health record (EHR) are first processed by an Event Splitter module that routes each note to condition-specific extraction pipelines spanning six irAE subtypes (pneumonitis, hepatitis, colitis, myocarditis, thyroiditis, and dermatitis), enabling parallel processing across organ systems. The pipeline proceeds through four sequential stages: (1) Detection — binary classification of irAE presence using self-consistency decoding (×3 runs) with judge adjudication; (2) Grading — severity assignment per CTCAE v5.0 criteria using self-consistency decoding (×3 runs) with consensus-based grade selection; (3) Attribution — immune checkpoint inhibitor (ICI) causality assessment via a single-agent causal reasoning step; and (4) Certainty — diagnostic confidence estimation via a single-agent certainty analysis step. Self-consistency is applied to the detection and grading stages, where classification ambiguity is highest, while attribution and certainty rely on single-pass inference given their more subjective, reasoning-intensive nature. The pipeline produces structured JSON output with supporting evidence traces for each identified event.

**Table 1:** Primary Performance Summary.

|  | Grade |  |  |  |  |  | Attribution |  |  | Certainty |  |  |
| --- | --- | --- | --- | --- | --- | --- | --- | --- | --- | --- | --- | --- |
|  | Binary F1 |  |  | Macro F1 |  |  | Binary F1 |  |  | Binary F1 |  |  |
| Condition | Current | Past | Agnostic | Current | Past | Agnostic | Current | Past | Agnostic | Current | Past | Agnostic |
| **Overall** | **0.92** | **0.91** | **0.93** | **0.66** | **0.76** | **0.75** | **0.92** | **0.94** | **0.94** | **0.93** | **0.94** | **0.95** |
| Thyroiditis | 0.94 | 0.97 | 1.00 | 0.72 | 0.82 | 0.91 | 0.87 | 1.00 | 0.95 | 0.87 | 1.00 | 0.95 |
| Hepatitis | 0.96 | 0.91 | 0.97 | 0.66 | 0.82 | 0.74 | 0.98 | 0.95 | 0.97 | 0.98 | 0.98 | 0.98 |
| Colitis | 0.87 | 0.95 | 0.90 | 0.51 | 0.78 | 0.70 | 0.93 | 0.92 | 0.96 | 0.91 | 0.88 | 0.92 |
| Pneumonitis | 0.96 | 0.93 | 0.94 | 0.67 | 0.74 | 0.67 | 0.88 | 0.96 | 0.95 | 0.96 | 0.95 | 0.97 |
| Myocarditis | 0.86 | 0.81 | 0.85 | 0.67 | 0.52 | 0.64 | 0.93 | 0.91 | 0.91 | 0.90 | 0.88 | 0.90 |

Retrospective performance of the irAE detection and grading pipeline across six immune-related adverse event subtypes. Current Grade Multi-class Macro F1: Macro-averaged F1 score across all 6 grade classes (0–5) for current conditions. For classes with no examples, F1 is set to 1.0. Macro-averaged metrics for binary classification (Grade 0 vs Grade 1+) for current conditions. These are the averages of metrics for both class 0 and class 1. Overall (Average Across Conditions): Simple unweighted average of each metric across all 6 conditions (Thyroiditis, Hepatitis, Colitis, Pneumonitis, Myocarditis, Dermatitis). Results with GPT-4.1-mini. Temporality settings: Current = current episode only; Past = historical mentions only; Agnostic = no temporal distinction.

Multi-class Common Terminology Criteria for Adverse Events (CTCAE) grade assignment showed lower performance with an overall macro F1 of 0.66 for current events. Performance varied by condition: dermatitis and thyroiditis achieved the highest performance (F1 = 0.73 and 0.72, respectively), while colitis had the lowest performance (F1 = 0.51). Intermediate grades (2–3) accounted for the majority of misclassifications. The system demonstrated stronger performance when severity was dichotomized at clinically relevant thresholds (grade <2 versus ≥2, and grade <3 versus ≥3), although performance varied according to irAE type (Figure 3).

**Figure 3.** Performance and efficiency of AI-assisted irAE grading. **a**,`Retrospective severity classification (Phase 1, n = 263 notes) across irAE types. Bars show F1 scores for exact grade (blue), grade <2 vs. ≥2 (orange), and grade <3 vs. ≥3 (grey).` **b**, Prospective validation (Phase 2, n = 884 notes) showing binary (blue) and macro (orange) F1 scores for irAE detection. **c**, Comparison of annotation time between manual (grey) and LLM-assisted (blue) review in the user effect study (Phase 3, n = 316 notes). Box plots display median and interquartile range (IQR); whiskers extend to 1.5×IQR; dots represent individual notes. * P < 0.001 (Mann–Whitney U test). **d**, Impact of AI assistance on efficiency and agreement metrics (Phase 3). Bar charts compare control (grey) vs. LLM-assisted (blue) conditions for time per note, accuracy, and inter-rater agreement (Cohen’s $\kappa$). Error bars denote 95% confidence intervals. ** P < 0.01; * P < 0.001.

The system reliably captured clinician-documented attribution assessments, with a binary F1 of 0.92 (current events) and 0.94 (past events). Certainty estimation remained high for binary classification (any stated certainty vs. none; F1 = 0.93–0.94).

The modest difference between temporality-agnostic and timepoint-specific detection (F1 = 0.93 vs. 0.92/0.91) indicates that residual errors were driven primarily by missed detections or incorrect grading rather than confusion between active and resolved events (Table 1).

Within the GPT-4.1-mini family, the single-inference variant (without self-consistency) reduced detection F1 from 0.92 to 0.78, while approximately halving inference cost ($0.011 vs. $0.021 per note), confirming the contribution of multi-run consensus to overall performance. Smaller models (4.1-nano, 4o-mini) and open-source alternatives consistently underperformed across detection and grading tasks (Appendix Table 1). Notably, a rule-based baseline using keyword-matching regular expressions^1^ achieved reasonable performance for binary detection (i.e., flagging whether an irAE was present or absent), performing comparably to or better than even some LLM-based methods. This finding underscores the utility of simple pattern-matching approaches for case identification; however, regular expressions are inherently limited to surface-level keyword matching and cannot extract the granular clinical attributes (such as severity grade, attribution, certainty, and supporting evidence spans) required for actionable irAE characterization.

### Phase 2: Prospective Silent Validation

To assess real-world generalizability, we deployed the system in silent, prospective mode for 3 months (May–July 2025), processing 884 clinical notes in near-real time. The system generated 31,824 label adjudications across grade, attribution, and certainty for both current and past timepoints. We found that detection performance remained robust, though attenuated relative to retrospective benchmarks.

Prospective performance was lower relative to retrospective benchmarks, as anticipated, due to temporal drift in documentation patterns and case mix (Table 2). In addition to the results in Table 2, detection F1 was 0.72 for current events and 0.79 for past events. Attribution extraction remained strong (F1 = 0.77), as did certainty classification (F1 = 0.80). At the note level, model predictions matched human review in 62% of cases for grade (549/884), 87% for attribution (771/884), and 83% for certainty (732/884).

**Table 2:**
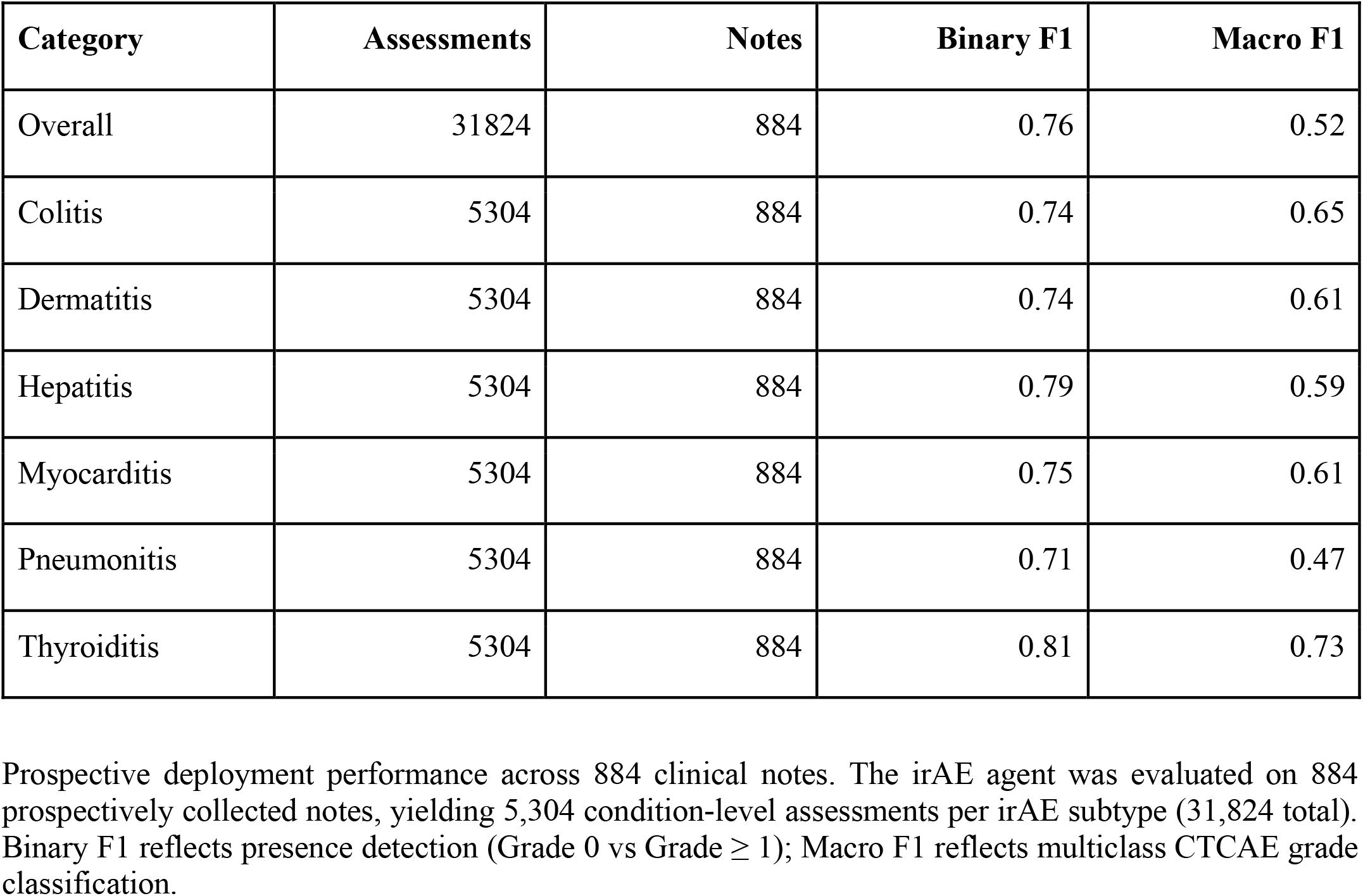
Prospective Performance (Overall and by Condition)

| Category | Assessments | Notes | Binary F1 | Macro F1 |
| --- | --- | --- | --- | --- |
| Overall | 31824 | 884 | 0.76 | 0.52 |
| Colitis | 5304 | 884 | 0.74 | 0.65 |
| Dermatitis | 5304 | 884 | 0.74 | 0.61 |
| Hepatitis | 5304 | 884 | 0.79 | 0.59 |
| Myocarditis | 5304 | 884 | 0.75 | 0.61 |
| Pneumonitis | 5304 | 884 | 0.71 | 0.47 |
| Thyroiditis | 5304 | 884 | 0.81 | 0.73 |

Prospective deployment performance across 884 clinical notes. The irAE agent was evaluated on 884 prospectively collected notes, yielding 5,304 condition-level assessments per irAE subtype (31,824 total). Binary F1 reflects presence detection (Grade 0 vs Grade $\geq 1$ ); Macro F1 reflects multiclass CTCAE grade classification.

### Phase 3: Randomized User Effect Study

To determine whether agentic assistance improves human curation of irAEs in practice, we conducted a randomized 2×2 crossover study comparing AI-assisted annotation to standard manual review among clinical research staff (Methods). We found that AI assistance substantially reduced annotation time, improved accuracy, and enhanced inter-annotator agreement. Twenty-one participants enrolled in the study (20 clinical research coordinators and 1 clinical research nurse), and 4 participants withdrew consent before starting annotations (all clinical research coordinators). All 17 remaining participants (16 clinical research coordinators, 1 clinical research nurse) completed the study (Appendix Table 1), yielding 316 evaluable timing observations for the primary note-level analysis. Twenty-four note-level observations across participants were excluded due to missing video recordings.

Table 3 shows results for the randomized user effect study. For our primary endpoint of annotation efficiency, AI assistance substantially reduced the time required to curate irAE labels from clinical notes. Median annotation time was 428 seconds (IQR: 253–671) with manual review versus 242 seconds (IQR: 160–385) with AI assistance—a 44% reduction. In the primary generalized estimating equations (GEE) analysis, which accounted for within-participant clustering, AI assistance was associated with a 40% decrease in annotation time (95% CI: 26–50%; P < 0.001). Neither note set nor method sequence significantly influenced efficiency in multivariable models, suggesting no substantial carryover effects.

**Table 3.** Time Efficiency, Accuracy, and Inter-Annotator Agreement (Control vs LLM)

| Section | Metric | Control | LLM-Assisted | Improvement |
| --- | --- | --- | --- | --- |
| **Timing** |  |  |  |  |
| Overall | Mean minutes/note | 7.80 (5.96–9.64) | 5.47 (3.57–7.38) | 2.32 (1.10–3.54)<br>**(p=0.004)** |
| **Accuracy (exact-match vs gold)** |  |  |  |  |
| Grade (overall) | % | 0.800 (0.764–0.841) | 0.843 (0.829–0.857) | 0.043 (-0.006–0.086) (p=0.791) |
| Grade (current) | % | 0.763 (0.718–0.814) | 0.809 (0.791–0.828) | 0.046 (-0.014–0.101) (p=0.607) |
| Attribution (overall) | % | 0.864 (0.837–0.894) | 0.928 (0.917–0.938) | 0.064 (0.032–0.092) **(p=<0.001)** |
| Attribution (current) | % | 0.841 (0.809–0.877) | 0.933 (0.924–0.942) | 0.092 (0.054–0.127) **(p=<0.001)** |
| Certainty (overall) | % | 0.798 (0.766–0.835) | 0.855 (0.847–0.862) | 0.056 (0.021–0.087) **(p=0.021)** |
| Certainty (current) | % | 0.764 (0.718–0.818) | 0.854 (0.842–0.866) | 0.090 (0.042–0.131) **(p=<0.001)** |
| **Inter-annotator Agreement (Cohen's kappa)** |  |  |  |  |
| Grade (overall) | % (kappa) | 0.444 (0.410–0.581) | 0.785 (0.744–0.825) | 0.341 (0.221–0.357) **(p=<0.001)** |
| Grade (current) | % (kappa) | 0.424 (0.381–0.556) | 0.825 (0.797–0.853) | 0.400 (0.266–0.446) **(p=<0.001)** |
| Attribution (overall) | % (kappa) | 0.404 (0.360–0.537) | 0.772 (0.729–0.816) | 0.369 (0.245–0.403) (**p=<0.001**) |
| Attribution (current) | % (kappa) | 0.357 (0.302–0.508) | 0.822 (0.787–0.858) | 0.465 (0.315–0.520) (**p=<0.001**) |
| Certainty (overall) | % (kappa) | 0.353 (0.252–0.470) | 0.758 (0.714–0.801) | 0.405 (0.292–0.501) (**p=<0.001**) |
| Certainty (current) | % (kappa) | 0.383 (0.289–0.508) | 0.793 (0.754–0.831) | 0.410 (0.272–0.517) (**p=0.002**) |
| *Timing p-value from two-sided paired sign test across participants. Accuracy p-values from paired sign test on within-participant accuracy differences (LLM – Control). IAA p-values from paired sign test across matched rater pairs; kappa is quadratic-weighted for grade and certainty, unweighted for attribution. 95% CIs are normal approximations.* |  |  |  |  |

Human–computer interaction study comparing unassisted (control) and LLM-assisted irAE annotation. (a) Time efficiency, accuracy, and inter-annotator agreement across grading, attribution, and certainty tasks. Timing is reported as mean minutes per note. Accuracy reflects exact-match agreement with a gold-standard reference. Inter-annotator agreement is reported as Cohen's kappa (quadratic-weighted for grade and certainty; unweighted for attribution),

For our secondary endpoint of performance, complete-match accuracy (requiring all six current grade labels to exactly match the gold standard) was modest overall, reflecting the stringency of this metric: 19.4% for manual annotation versus 24.9% with AI assistance (Table 3). In GEE analysis, AI assistance increased the odds of complete accuracy by 45% (OR = 1.45; 95% CI: 1.01–2.09; P = 0.045). This effect remained significant after adjustment for note set and sequence.

Further, for inter-annotator agreement AI assistance markedly improved consistency across annotators. For current grade labels, Krippendorff’s alpha increased from 0.22 (95% bootstrap CI: 0.16–0.30) to 0.85 (95% CI: 0.79–0.89) in note set A, and from 0.51 (95% CI: 0.50–0.51) to 0.82 (95% CI: 0.76–0.87) in note set B. The magnitude of improvement was substantial in both sets (Δα > 0.30), indicating that AI assistance reduced between-annotator variability and promoted standardization. Exploratory analyses of attribution and certainty labels showed concordant improvements: for attribution, Krippendorff’s α increased from 0.22 to 0.86 (note set A) and from 0.42 to 0.75 (note set B); for certainty, from 0.18 to 0.84 and from 0.44 to 0.74, respectively.

Participants’ perceptions mirrored these objective gains (Appendix Table 2). Mean System Usability Scale scores increased from 35.3 (manual) to 52.1 (AI-assisted). On 5-point Likert items from pre- and post-study questionnaires (Appendix 5b, 5c), self-reported efficiency increased from 2.5 to 3.9, confidence in labeling accuracy from 3.1 to 3.8, and belief that AI can assist complex clinical interpretation from 2.3 to 3.0. Following the study, 88% of participants preferred the AI-assisted workflow to manual review.

## Discussion

The present study explores an agentic LLM system for the extraction of clinically salient attributes of irAEs from routine oncology documentation with performance that translates from retrospective benchmarking to prospective, real-world use and improved adverse event data quality. Across six high-priority irAEs, the system achieved strong note-level detection and clinician-stated attribution performance. Multi-class CTCAE grading was promising, with degradation at intermediate grades. Performance declined during a three-month prospective silent deployment (F1 = 0.72–0.79), underscoring the gap between retrospective evaluation and changing real-world conditions. In a randomized user effect study, agentic-assistance reduced annotation time by 40%, improved accuracy, and improved inter-annotator reliability. Together, these results suggest that agentic LLM systems, coupled to human verification and evidence highlighting, could be further explored to shift irAE ascertainment from a labor-intensive retrospective review to scalable, efficient automated extraction with a potential to improve consistency and quality.

Prior approaches emphasized patient- or visit-level irAE classification, retrospectively and without the fine-grained attributes required for actionability and causal effects analysis (temporality, grade, and attribution).^21,25,26,28^ By decomposing the task into specialized agents for temporality, grading, attribution, and certainty with explicit extraction of supporting spans, our system overcomes known challenges in textual causality and nuance.^29,32^ The self-consistency mechanism, in which a judge agent reconciled three parallel model runs for a stage, contributed materially to detection performance (macro F1 +0.14 relative to the single-inference variant), consistent with the value of ensembling and cross-examination to reduce output variance.^33,34^ Temporal misassignment, the system labelling an irAE as currently active when it was historical, or vice versa, was small (|ΔF1| < 0.05 across conditions). A cross-temporal analysis that credited a prediction if the irAE was detected in either timeframe yielded only modest gains (binary F1 = 0.93), confirming that residual errors are primarily detection- or grading-related rather than due to timeframe confusion.

These results carry direct clinical and pharmacovigilance implications. Reliable, note-level extraction of irAE attributes addresses a persistent blind spot in structured EHR fields and claims data, where recall is limited even for hospitalization-associated events. The ability to surface candidate events with attributed causality and provisional grade shortly after documentation can shorten time-to-recognition for life-threatening irAEs (e.g., myocarditis, pneumonitis) and standardize ascertainment for research registries and safety reporting.

Multi-class CTCAE grading remains the most challenging subtask. Two considerations temper this result. First, ambiguity at intermediate grades often reflects documentation incompleteness (e.g., missing quantitative stool counts for grading of colitis, absent laboratory trend context for grading of hepatitis) rather than model error per se; such ambiguity similarly burdens human reviewers. Second, thresholds can be adapted to clinically meaningful bins (e.g., none/low vs ≥ Grade 3), which may improve effective performance for triage and reporting while preserving granular outputs for research. Incorporating structured signals (laboratories, medications such as corticosteroid initiation, imaging impressions) and temporal aggregation across notes are promising future strategies to disambiguate intermediate grades without sacrificing interpretability.

The transition from retrospective benchmarking to prospective deployment revealed expected but instructive performance attenuation. The system was optimized on a curated retrospective corpus of several hundred notes, then deployed against a continuous stream of real-world documentation generated years later on different EHR templates, encompassing a broader range of clinical presentations, irAE subtypes, symptom descriptions, and documentation styles than were represented in the development set. This degradation, therefore, likely reflects the combined effects of distributional shift in case mix and clinical terminology, evolving EHR templates and note structures, and variation in documentation practices across clinician cohorts, rather than fundamental model limitations. The prospective phase thus underscores a critical operational requirement: deployed clinical AI systems require continuous monitoring and periodic recalibration,^35^ not one-time validation. Attribution and certainty extraction proved more robust to drift than severity grading, consistent with the relative stability of causal language conventions versus the context-dependent nature of quantitative symptom documentation.

The randomized user study provides, to our knowledge, the first controlled evidence that agentic AI assistance improves adverse event curation when deployed with human oversight. Three findings merit emphasis. First, efficiency gains were substantial and exceeded our pre-specified minimally important difference, suggesting that AI-prepopulated labels shift annotator effort from generation to verification—a cognitively less demanding task. Second, there was a striking effect on inter-annotator consistency: manual annotation produced only fair agreement, whereas AI-assisted annotation achieved near-excellent reliability. This convergence effect likely arises because annotators anchor on model suggestions, reducing idiosyncratic interpretation of ambiguous documentation. For pharmacovigilance and registry curation, where consistency across reviewers is as important as individual accuracy, this standardization benefit may prove as valuable as time savings. However, the same anchoring mechanism raises the risk of automation complacency, whereby reviewers accept model outputs without sufficient independent scrutiny, potentially introducing systematic bias. Our study was not designed to disentangle consistency gains from complacency effects, and future work should explicitly test whether anchoring on AI suggestions compromises the detection of model errors, particularly for subtle or atypical presentations. Third, participants’ qualitative feedback highlighted the value of evidence highlighting, the extraction and display of specific text spans from the clinical note (e.g., the phrases describing respiratory symptoms that supported a pneumonitis determination) alongside each model output. By linking each extracted label to its source evidence within the note, this feature enabled reviewers to more easily verify model conclusions against the original documentation directly, rather than re-reading entire notes, and served as a practical enabler of efficient human-AI collaboration.

These findings are consistent with a growing body of work suggesting that AI assistance can improve throughput in clinical documentation and annotation tasks, including ambient AI scribes that reduce documentation time by up to 30 minutes per clinician per day,^36^ and LLM-assisted radiology reporting that accelerates report generation while maintaining diagnostic accuracy.^37^ The 88% preference for the AI-assisted workflow, coupled with improvements in perceived efficiency and confidence, suggests that such systems can achieve the often-elusive goal of user acceptance in clinical informatics.

Questions of generalizability, governance, and cost warrant consideration. Interestingly, the architecture generalized across six irAEs with consistent detection performance, suggesting portability to a broader spectrum of adverse events. Nevertheless, this is a single-health-system study with local note templates and vernacular; external validation across institutions, EHRs, and oncology subspecialties is necessary. A practical barrier to adoption is cost and infrastructure. Agentic pipelines require multiple model calls per note, and our system currently relies on a proprietary model (GPT-4.1-mini) accessed via a commercial API, which incurs per-token inference costs that may be prohibitive for small and medium-sized institutions lacking enterprise API agreements or dedicated compute infrastructure. In the present study, the best-performing agentic configuration cost $0.021 per note (~$18.50 for the 884-note prospective corpus), while open-source alternatives running on institutional GPUs eliminated API costs entirely, albeit with lower performance (Appendix Table 1). Although our results with a mid-sized model suggest that performance– cost trade-offs can be managed at scale, broader adoption will likely require validating open-source or locally deployable LLMs that can run behind institutional firewalls under HIPAA BAAs, reducing both cost and reliance on external vendors. Our preliminary benchmarking of open-source alternatives showed lower performance with current prompting strategies (Appendix Table 1), but rapid improvements in open-weight models make this a tractable near-term goal. Robust monitoring for data drift will also be essential as indications and combination regimens expand.^38^

Several limitations should be acknowledged. We focused on six irAEs representing a subset of the broader CTCAE v5.0 ontology; expansion to rarer toxicities will require targeted evaluation. Our system was developed and validated using a single proprietary model (GPT-4.1-mini), and performance may not transfer directly to other LLMs; systematic evaluation across open-source and locally deployable models is an important next step for broader accessibility. Ground-truth labels, though adjudicated, remain constrained by documentation quality and the inherent subjectivity of grading and attribution. The prospective deployment did not assess patient outcomes (e.g., time to steroid initiation, hospitalization, treatment interruption). The preparatory field study was observational; causal effects on efficiency and agreement were isolated in the randomized study (Phase 3). While we only studied irAEs, we believe that the results will be informative for other cancer AE’s and adverse events more broadly. In particular, the human-computer interaction results may be transferable for systems with similar performance and interface features.

Looking ahead, these findings carry both methodological and practical implications. Methodologically, agentic decomposition with self-consistency and explicit evidence extraction yielded accurate, auditable outputs suitable for clinical workflows. Practically, staging validation from retrospective testing to silent prospective monitoring and embedded human collaboration provides a template for closing the “deployment gap” in clinical AI. Given the expanding survivor population exposed to ICIs and diffusion of care into non-oncology settings, scalable, auditable assistance for irAE detection is poised to become a core informatics capability for oncology services and pharmacovigilance programs.

## Conclusions

In summary, an agentic LLM system and associated interactive interface, validated across retrospective and prospective settings, can extract real-time, clinically detailed information about the presence, severity, and attribution of irAEs. Agentic assistance improved the efficiency, performance, and consistency of irAE curation when used with a human-in-the-loop. With external validation and outcome-focused trials, such systems could enhance and accelerate real-time safety monitoring, trial curation, and high-reliability oncology care.

## Methods

### Study Design

This multi-phase study evaluated an agentic LLM system for detecting and extracting irAEs from clinical documentation, progressing systematically from retrospective analysis to real-world deployment (Figure 1). We first developed and retrospectively evaluated a note-level agentic irAE extraction system. We then conducted real-world validation studies to (1) validate prospective performance when integrated within an EHR for real-time clinical monitoring, and (2) quantify the impact of human-agentic collaboration on irAE curation efficiency and quality via a randomized user effect study. A preparatory field study was conducted to inform interface design, metric selection, and power calculations. The study protocol received ethical approval from the Institutional Review Boards of Mass General Brigham and Dana-Farber Cancer Institute (MGB 2022P002195, DF/HCC 24-735, DF/HCC 20-328). A waiver of informed consent was granted for the retrospective components, as the research involved minimal risk and did not alter any clinical care. Electronic informed consent was obtained for the human-computer interaction studies.

### Phase 1: Note-Level irAE System Development and Evaluation

For the data source and study population, we assembled a retrospective cohort of patients treated with immune checkpoint inhibitors at an academic medical center in Boston, MA between 2015 and 2024. Inclusion criteria were: (1) age ≥18 years, (2) treatment with one or more of the following FDA-approved ICIs: *tremelimumab, relatlimab, retifanlimab, pembrolizumab, nivolumab, atezolizumab, durvalumab, avelumab, cemiplimab, toripalimab, dostarlimab*, and (3) presence of at least one progress note or visit note in the clinical system. Demographic information of included participants across phases are included in Appendix 2.

To establish the gold-standard annotation procedure, annotation guidelines were developed to support manual ground-truth labeling of the presence of current and past irAEs and their key clinical attributes (Appendix 3). We focused on 6 irAEs, which were selected to capture a range of common, rare, severe, and chronic events: pneumonitis, hepatitis, myocarditis, dermatitis, thyroiditis, and colitis. Each irAE was mapped to a set of National Cancer Institute (NCI) CTCAE terms following the definitions used in the Alliance for Clinical Trial in Oncology irAE biobank (NCT04242095, Appendix 3). For each irAE type and temporality (current vs. past) combination, we labeled its binary presence or absence, CTCAE grade, attribution, and certainty as described in a note. The clinical attributes are defined as follows: **(1)** Severity: Severity grading followed the National Cancer Institute Common Terminology Criteria for Adverse Events (CTCAE) version 5.0, a standardized system commonly used for regulatory reporting. Values: Grade 1, 2, 3, 4, 5. **(2)** Attribution: Attribution captured the clinician’s explicitly documented assessment of causality between the adverse event and ICI therapy, recognizing that many symptoms in cancer patients have multiple potential etiologies. Attributes: Positive, Negative.

**(3)** Certainty: Certainty reflected the degree of diagnostic confidence expressed in the documentation, guided in part by the Naranjo scale but adjusted for note-level, text-based determination.^39^ Attributes: None, Unlikely, Possible, Probable, Definite.

Pilot rounds of 10-20 notes were dual-annotated by a physician and clinical research coordinator with 4 years of oncology data abstraction experience, and annotation guidelines were iteratively updated for clarity and consistency until inter-annotator agreement exceeded 0.70 for each irAE type. All discrepancies were tracked and resolved through consensus discussion. Cases requiring adjudication were reviewed by an attending oncologist. The open-source Label Studio Community Platform v1.15 (HumanSignal, San Francisco, US), locally hosted behind our institution’s firewall, was used as our annotation platform.

A sample of notes from 100 patient encounters, forming a total of 289 retrospective patient notes, were annotated for retrospective evaluation (Appendix 2). These notes were divided into a development set of 26 notes, all from pneumonitis-positive patients, and a test set of 263 notes, used to evaluate the final developed system.

For the development of the agentic LLM architecture, OpenAI application programming interface (API)-based models were accessed using our institutional HIPAA-compliant application programming inference access points. Open-source models were downloaded locally, behind our institutional firewall, to a dedicated server equipped with two NVIDIA RTX 6000 Ada GPUs.

The architecture employed an agentic design pattern, decomposing the complex task of irAE extraction into specialized subtasks handled by distinct model instances (Figure 2). A preprocessing agent first extracted and formatted relevant clinical text, handling the varied structures and formats present in EHR documentation. Subsequently, specialized extraction agents focused on specific attributes: one agent determined temporal status by analyzing verb tenses and temporal markers, another mapped clinical descriptions to CTCAE grades, a third assessed attribution by identifying causal language and clinical reasoning, and a fourth evaluated the certainty of diagnosis based on hedging language and diagnostic workup discussions. We refer to this as the “default” agentic architecture hereafter. Prompt engineering was optimized on the development set, and only for pneumonitis, to assess the generalizability of our approach to other irAE types.

Given the potential variability in LLM outputs and the risk of hallucination, we also evaluated adding a self-consistency mechanism.^33,34,40^ Here, each note was processed through three parallel inference runs with controlled randomness. A final judge agent then synthesized these parallel outputs, implementing a majority-vote mechanism for discrete categories and selecting the most coherent extraction when unanimity was absent.

Finally, based on preparatory field study work (Appendix 4), the models were designed to extract “string-matched” snippets directly from clinical notes, as well as a brief rationale for the determinations, to support human interaction in real-world downstream tasks. The snippets serve as supporting evidence for the determinations made at each stage of the agentic workflow. This functionality was then integrated into the user interface as features to aid human reviewers in their annotation tasks.

As comparative baseline methods, to contextualize the performance of our agentic system, we implemented several baseline approaches representing different points on the complexity-performance spectrum. These included rule-based extraction using regular expressions previously used in our institution to identify patients admitted for irAEs^41^; and zero-shot prompting. We compared open-source (Qwen-3-4B-Instruct, Qwen3-30B-A3B-Instruct-2507, Gemma-3-27B, GPT-OSS-20B) and proprietary OpenAI models (GPT-4.1-mini, GPT-4.1-nano, gpt-4o-mini) with medium reasoning effort on our institutional HIPAA-secure OpenAI API endpoint. Model selection was guided by the goal of representing a range of model sizes deployable on single- and dual-GPU institutional hardware for open-source candidates, alongside cost-constrained proprietary models available under Azure HIPAA-compliant agreements.

Precision, recall, and macro-averaged F1 scores were calculated overall and stratified by irAE type across all available classes for a given label. The inference cost per note was also calculated for all variants.

### Phase 2: Prospective Silent Validation

Following retrospective analysis, we implemented the best setting of our system in a silent, prospective, real-time clinical environment to assess performance under operational conditions and to assess generalizability to evolving clinical language and documentation practices. The system was integrated with our institution’s clinical data warehouse. Details of the integration infrastructure have been previously reported.^38^ New Physician, Nursing, and Physician Assistant notes documented on patients treated with an ICI within the preceding year were automatically identified each morning, including progress notes, radiology reports, consultation reports, emergency department records, and discharge summaries. Notes meeting criteria were immediately processed through the agentic system. The first 102 notes were dual-annotated by a board-certified oncologist and a trained research assistant with experience as a medical scribe and clinical research, achieving an inter-annotator agreement across irAE type and temporality (κ = 0.91 for binary detection, κ = 0.88 for grade). The remaining notes were single-annotated by the research assistant. This prospective phase lasted for three months (May-July 2025), during which we processed 884 real-time clinical notes. Agreement between prospective model predictions and human determinations was quantified using agreement percentages and override rates for each label and across all labels.

### Phase 3: Randomized User Effect Study

To quantify the clinical impact of agentic assistance on irAE curation, we conducted a 2×2 randomized crossover study comparing AI-assisted annotation to standard manual review (Appendix Figure 2). This design was informed by a preparatory field study that informed the interface design, characterized learning curves, established baseline inter-rater agreement, and identified annotation efficiency as the primary driver of real-world workflow burden (Appendix 4). This study was approved by the Institutional Review Board (DF/HCC 20-328).

We enrolled clinical research coordinators and clinical research nurses from our department; there were no exclusion criteria. Following informed consent, participants completed a one-hour standardized training session with annotated examples (Appendix 5a) and an unobserved practice period using the annotation interface (Figure 4). Participants were then randomized to one of four sequences in a crossover design that counterbalanced both intervention order (AI-assisted vs. manual spreadsheet) and note set (Set A vs. Set B). This structure was chosen to mitigate carryover effects from either the annotation method or note content, and enabling difference-in-differences estimation of treatment effects.

**Figure 4.** Screenshot of the Labelling interface used for chart curation with the IrAE-Agent. The annotation interface takes in the outputs from the IraE-Agent and allows users to label the notes across conditions while highlighting extracted evidence in the note pane. All timings and model agreement metrics are automatically stored.

Clinical notes were drawn from the Phase 1 retrospective corpus. Sets A and B were matched on note length and clinical complexity and number of irAE descriptions through manual review to ensure comparability across arms.

Based on pilot data from the preparatory field study (Appendix 4), we estimated a clinically meaningful efficiency gain of 450 seconds per note (SD = 192 seconds). After inflating the sample size to account for the within-participant correlation of 0.05, 15 participants completing 20 notes each (300 total notes) would provide 84.7% power to detect a standardized effect size of −0.43 (comparing AI assistance to manual) with two-sided α = 0.10. We targeted enrollment of 17–20 participants to account for potential dropouts and missing observations.

The primary endpoint was annotation efficiency, measured as time in seconds from note opening to final submission, captured via screen recording. Secondary endpoints were: (1) accuracy, defined as exact match with the gold standard for all current irAE grades within a note—a stringent metric requiring perfect concordance across all six conditions; and (2) inter-annotator agreement, assessed using Krippendorff’s alpha for ordinal grade labels.

For the primary efficiency endpoint, we used generalized estimating equations (GEE) with an exchangeable correlation structure to account for within-participant clustering and estimated the reduction in annotation time attributable to AI assistance. Sensitivity analyses examined log-transformed times, adjusting for note set and sequence effects.

For the accuracy endpoint, we used GEE with binomial outcomes to estimate the odds ratio of exact gold-standard match comparing AI-assisted to manual annotation, with robust sandwich variance estimators. Additional exploratory analyses relaxed the stringent complete-match criterion to examine accuracy for individual label types (grade, attribution, certainty) and by temporality. Further, inter-annotator reliability was assessed separately within each note set using Krippendorff’s alpha for current grade labels. Bootstrap resampling (10,000 iterations, resampling participants with replacement) generated 95% confidence intervals for each method. Pre- and post-study questionnaires captured baseline AI attitudes, the System Usability Scale, and perceived efficiency (Appendix 5b and 5c). Participants were compensated $50/hour.^42^

## Code and Data Availability

The code and prompts will be available at: https://github.com/BittermanLab/AEGIS following acceptance. Raw data has not been deidentified and is not available for public distribution. Derived results in aggregate are available in the repository.

## Acknowledgements

The authors acknowledge financial support from the National Institutes of Health National Cancer Institute (U54CA274516-01A1 [J.G., D.S.B, D.E.K., G.K.S.], R01CA294033-01 [J.G., P.F.D., B.Y., M.H.., J.L.W, M.J.H., E.S., D.E.K., G.K.S., D.S.B], 2U24CA248010 [D.S.B., G.K.S., H.H., J.L.W.]), U01209414 to DCC and RHM), the American Cancer Society and American Society for Radiation Oncology, ASTRO-CSDG-24-1244514-01-CTPS Grant DOI #: https://doi.org/10.53354/ACS.ASTRO-CSDG-24-1244514-01-CTPS.pc.gr.222210[D.S.B.], a Patient-Centered Outcomes Research Institute (PCORI) Project Program Award (ME-2024C2-37484) [D.S.B.], and the Woods Foundation [D.S.B.]. All statements in this report, including its findings and conclusions, are solely those of the authors and do not necessarily represent the views of the Patient-Centered Outcomes Research Institute (PCORI), its Board of Governors or Methodology Committee.

## Prior presentations

None.

## Author Contributions

J.Ga., K.C.K., and D.S.B. conceptualized the study. J.Ga. and K.C.K. performed the interviews. J.Ga. wrote the first draft of the manuscript, and J.Ga., S.C, K.C.K., and D.S.B. critically reviewed and edited subsequent versions. M.B., A.C., S.C., P.F.D., S.D., J.Gu., M.J.H., E.H.K., D.E.K., H.H., J.W.L.A., A.B.L., R.H.M., R.G.M., T.L.N., G.K.S., E.S., B.C.S., U.T., and J.L.W. contributed to study design, interpretation of findings, and critical review and revision of the manuscript.

## Footnotes

1 A fully optimized/customized regular expression that includes 779 terms expanded from our physician provided lists.

