## Supplementary figures and images for "An agentic AI system enhances clinical detection of immunotherapy toxicities: a multi-phase validation study"

### Appendix Figure 1

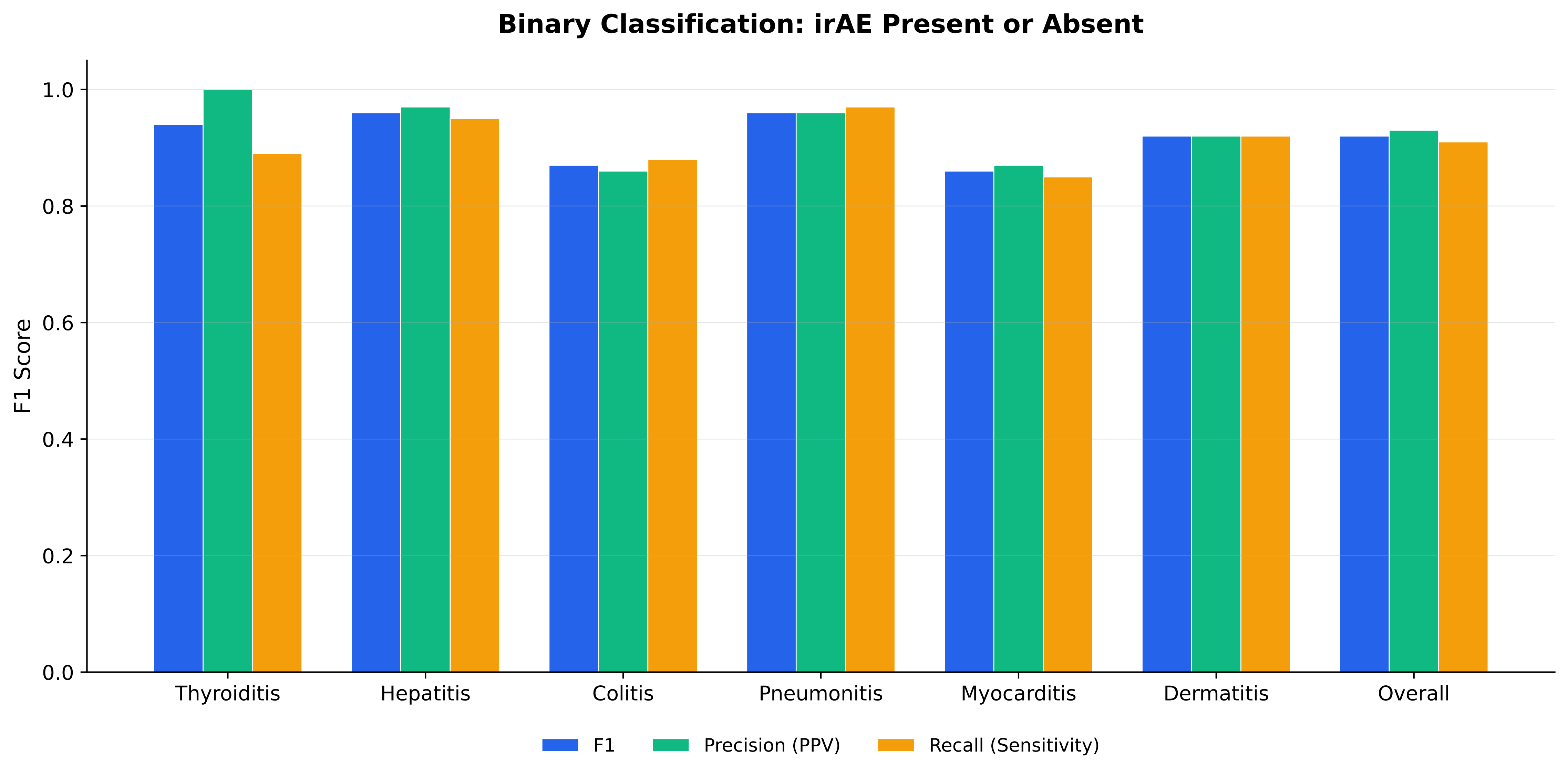

### Figure 1

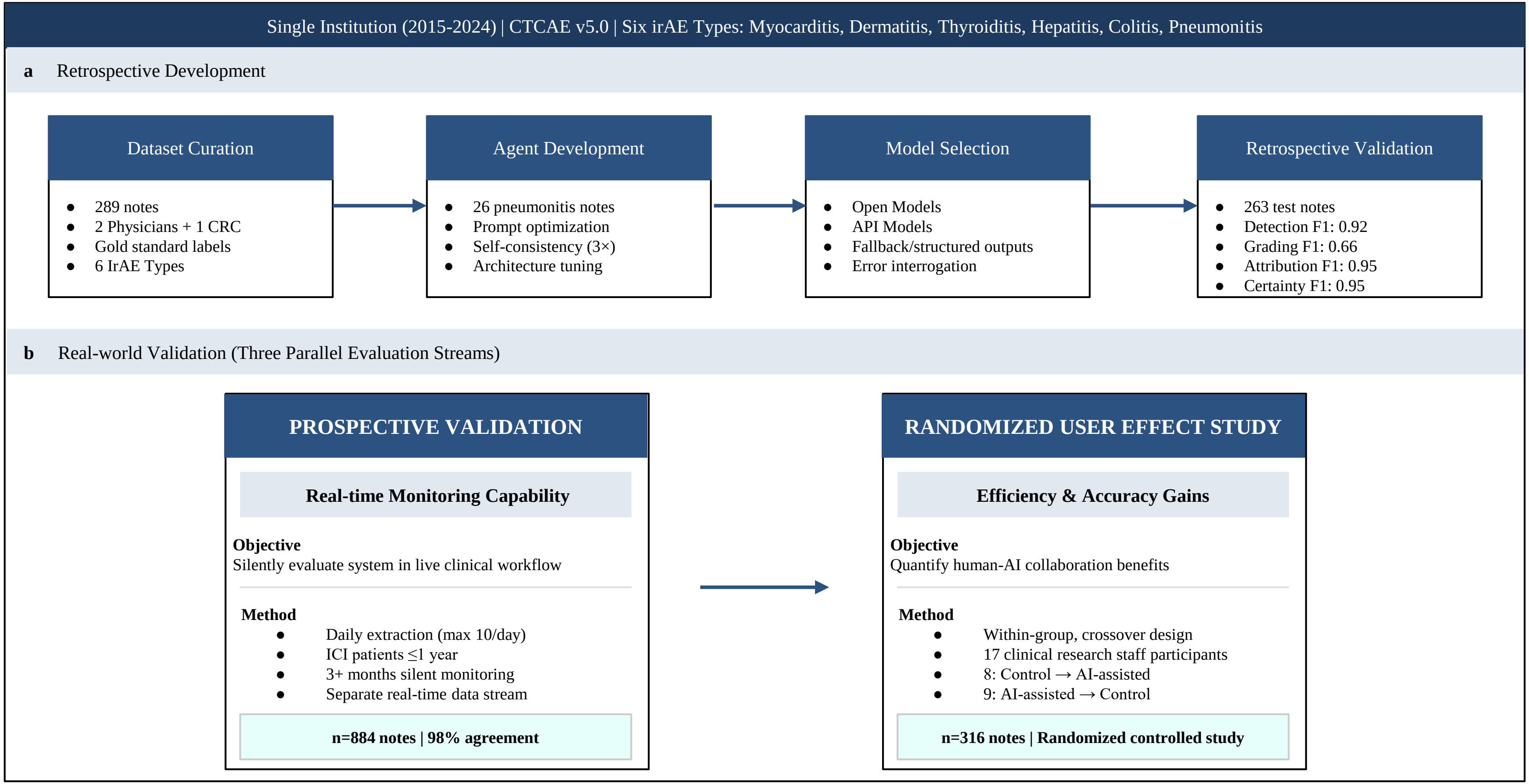

### Figure 2

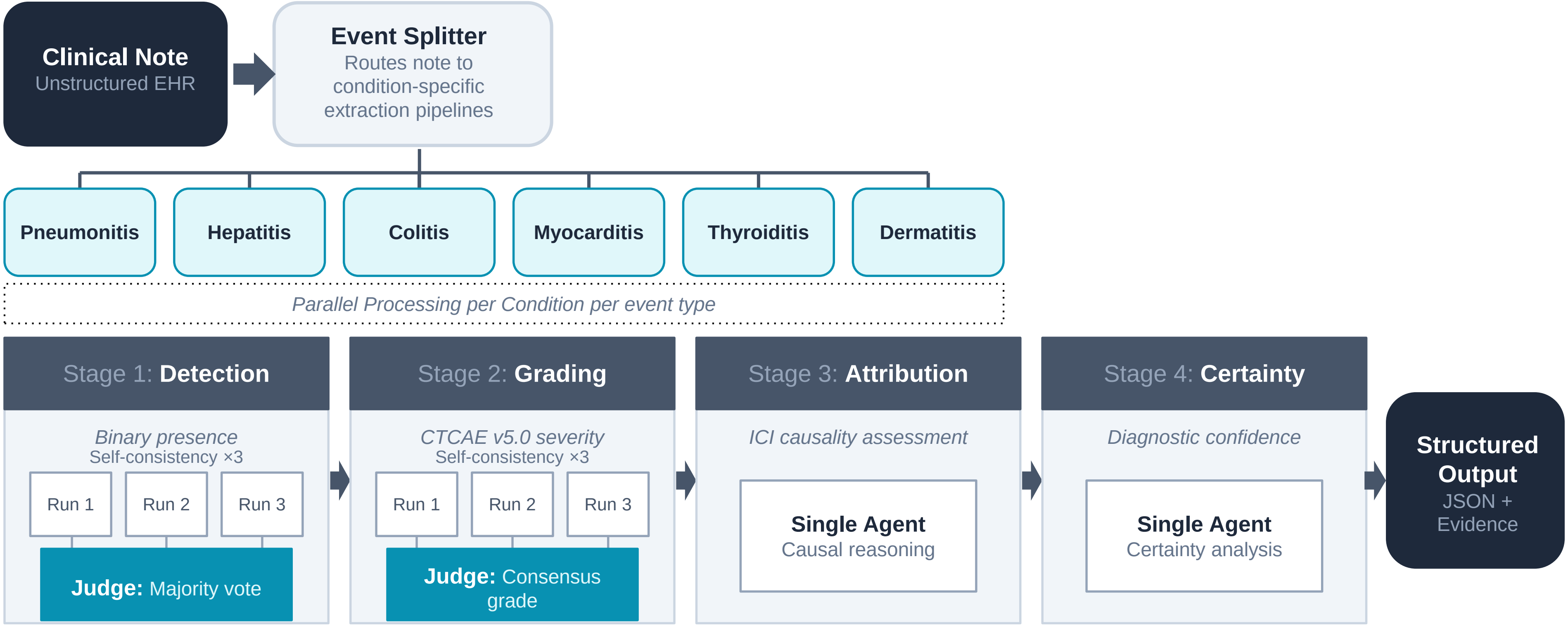

### Figure 3

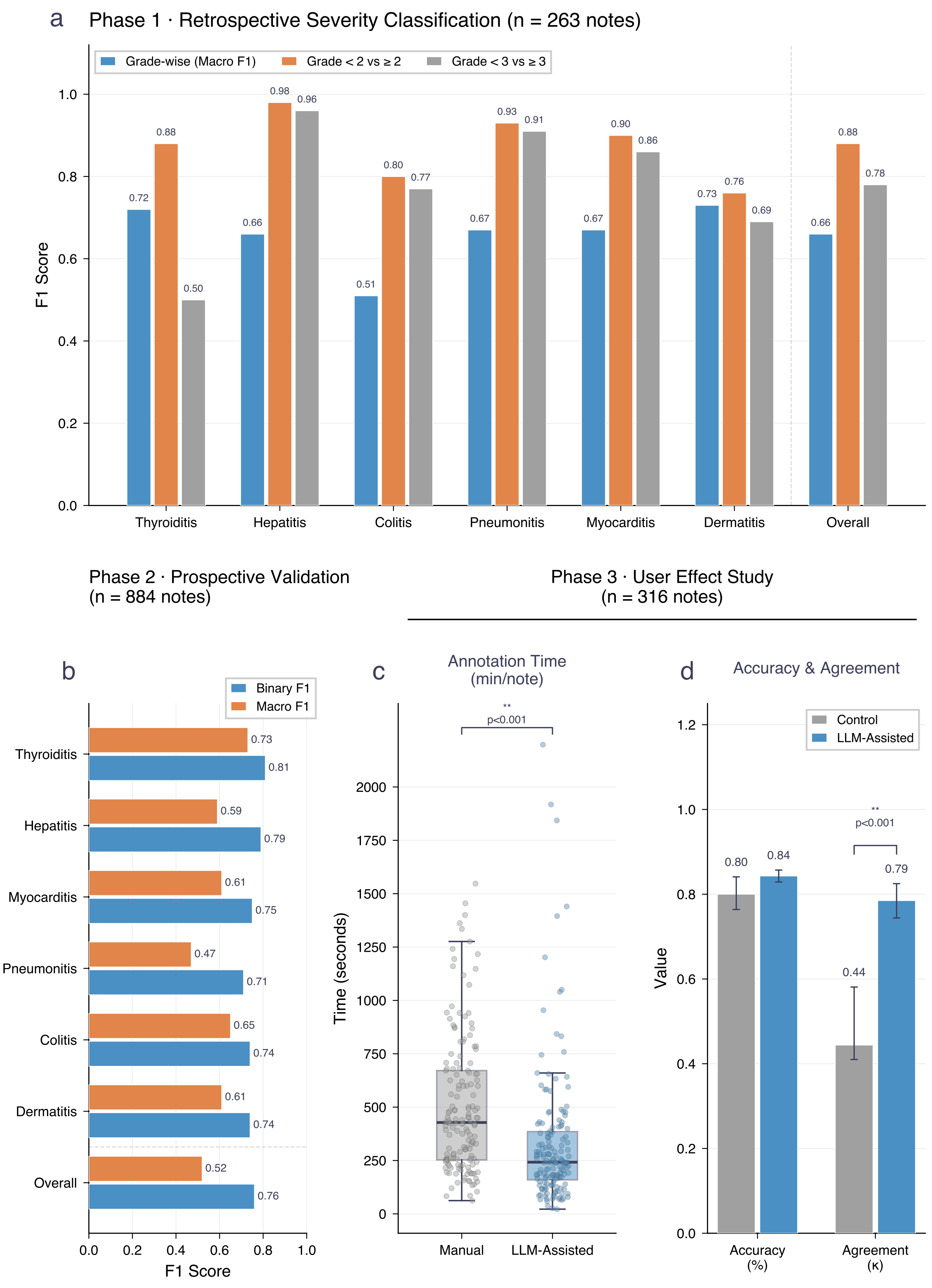
