## Supplementary material for "An agentic AI system enhances clinical detection of immunotherapy toxicities: a multi-phase validation study": Figure 4

### Evidence Clarity Demo - Labeling Interface

Back to Home

**Demo Mode:** This is a demonstration of the Evidence Clarity labeling interface. No real data is being used, and no changes are being saved.

Note 1/10 — 3 + Prev Next

Search in note...

ONCOLOGY CONSULTATION NOTE

Date: January 15, 2024
Patient: JOHNSON, ROBERT
MRN: DEMO-PT-001
DOB: 05/12/1958 (65 y.o.)
Provider: Dr. Emily Roberts, Oncology
Department: Hematology/Oncology

REASON FOR CONSULTATION:
Evaluate worsening respiratory symptoms in patient on pembrolizumab for metastatic NSCLC.

HISTORY OF PRESENT ILLNESS:
Mr. Johnson is a 65-year-old male with stage IV NSCLC, currently on cycle 8 of pembrolizumab monotherapy. He was last seen 3 weeks ago and was doing well. He now presents with progressive dyspnea over the past 10 days, associated with dry cough and decreased exercise tolerance. He reports difficulty climbing one flight of stairs without stopping to catch his breath. Denies fever, chest pain, or hemoptysis. No sick contacts or recent travel.

PAST MEDICAL HISTORY:
- Stage IV NSCLC with brain metastases, diagnosed 10 months ago
- Former smoker (40 pack-year history, quit at diagnosis)
- COPD, mild
- Hypertension
- Type 2 diabetes mellitus

MEDICATIONS:
- Pembrolizumab 200mg IV q3 weeks (last dose 12/28/2023)
- Metformin 1000mg BID
- Lisinopril 20mg daily
- Albuterol inhaler PRN

PHYSICAL EXAMINATION:
VS: BP 142/88, HR 96, RR 22, T 98.4°F, O2 Sat 91% on room air, 95% on 2L NC
General: Mild respiratory distress at rest
Lungs: Bilateral fine crackles, more prominent at bases. No wheezing.

Patient: DEMO-PT-001 Note: ONCO-001 Dept: Hematology/Oncology Jan 15, 2024 11:30

#### Labeling

Guidelines

Pneumonitis | Myocarditis | Colitis | Thyroiditis | Hepatitis | Dermatitis

##### User Overview

**Pneumonitis Overview**

**Pneumonitis Summary**

Pneumonitis was detected in the current context with grade 3 severity, indicating severe inflammation of lung tissue likely due to immunotherapy. Evidence includes CT findings showing patchy infiltrates and clinical symptoms of dry cough and shortness of breath.

##### Model Predictions

Current: G 2 C 4 — Use Current — Add Current

Evidence for Pneumonitis (6)

Relevant
Pembrolizumab-induced pneumonitis, Grade 2 - Clinical presentation and imaging findings consistent with imm...

Relevant
Metastatic NSCLC - Stable on recent imaging, no evidence of progression - Immunotherapy on hold due to pne...

Relevant
Heart: Regular rate and rhythm, no murmurs Extremities: No cyanosis or edema DIAGNOSTIC STUDIES: - CT ch...

Relevant

1/6

##### Label: Pneumonitis

Temporality: Current | Past

Grade: 0 | 1 | 2 | 3 | 4 | 5

Attribution: False | True

Certainty: 0 | 1 | 2 | 3 | 4

(None)
