## Appendix Figure 2 for "An agentic AI system enhances clinical detection of immunotherapy toxicities: a multi-phase validation study"

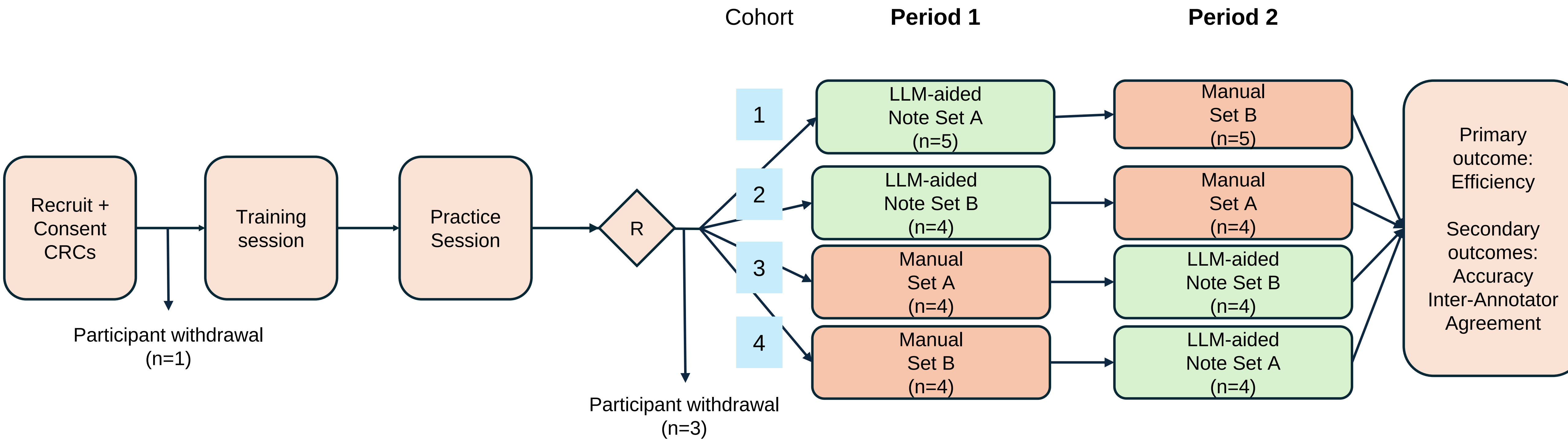


**Appendix Figure 2.** Randomized crossover study design. Participants completed training and unobserved practice sessions before randomization (R) into one of four cohorts. The 2×2 crossover design counterbalanced both annotation method (LLM-aided interface vs. manual spreadsheet) and clinical note set (Set A vs. Set B) across two periods to mitigate carryover effects. Primary endpoint: annotation efficiency. Secondary endpoints: accuracy against gold standard and inter-annotator agreement (κ).
