## Appendix Table 1 for "An agentic AI system enhances clinical detection of immunotherapy toxicities: a multi-phase validation study"

| Graph Type | Model | Variant | Overall Binary F1 | Overall Macro F1 |
|---|---|---|---|---|
| openai_agent | 4.1-mini | default | 0.92 | 0.66 |
| openai_zeroshot | ollama-gpt-oss-20B | default | 0.86 | 0.59 |
| openai_zeroshot | Gemma-3-27B | default | 0.84 | 0.53 |
| openai_zeroshot | 4.1-mini | default | 0.83 | 0.52 |
| openai_zeroshot | Qwen3-30B-A3B-Instruct-2507 | default | 0.83 | 0.62 |
| openai_agent | 4o-mini | ablation_no_judge | 0.82 | 0.65 |
| regex | default | default | 0.81 | 0.43 |
| openai_agent | 4o-mini | ablation_single | 0.79 | 0.64 |
| openai_agent | 4o-mini | default | 0.79 | 0.47 |
| openai_zeroshot | Qwen3-30B-A3B-Instruct-2507 | default | 0.78 | 0.51 |
| openai_agent | 4.1-mini | ablation_single | 0.78 | 0.64 |
| openai_agent | 4.1-nano | default | 0.78 | 0.52 |
| openai_agent | Qwen3-30B-A3B-Instruct-2507 | default | 0.78 | 0.59 |
| openai_agent | 4.1-nano | ablation_single | 0.77 | 0.53 |
| openai_zeroshot | Qwen-3-4B-Instruct | default | 0.77 | 0.52 |
| openai_agent | Qwen-3-4B-Instruct | default | 0.72 | 0.55 |
| openai_zeroshot | 4.1-nano | default | 0.71 | 0.45 |
| openai_agent | ollama-gpt-oss-20B | default | 0.70 | 0.53 |
| openai_agent | Gemma-3-27B | default | 0.48 | 0.38 |

| Thyroiditis Binary F1 | Thyroiditis Macro F1 | Hepatitis Binary F1 | Hepatitis Macro F1 | Colitis Binary F1 | Colitis Macro F1 | Pneumonitis Binary F1 |
|---|---|---|---|---|---|---|
| 0.94 | 0.72 | 0.96 | 0.66 | 0.87 | 0.51 | 0.96 |
| 0.83 | 0.64 | 0.91 | 0.50 | 0.84 | 0.43 | 0.94 |
| 0.81 | 0.76 | 0.91 | 0.64 | 0.93 | 0.40 | 0.88 |
| 0.81 | 0.68 | 0.86 | 0.52 | 0.85 | 0.43 | 0.81 |
| 0.80 | 0.79 | 0.91 | 0.66 | 0.80 | 0.43 | 0.90 |
| 0.68 | 0.74 | 0.68 | 0.49 | 0.94 | 0.81 | 0.84 |
| 0.81 | 0.66 | 0.89 | 0.33 | 0.86 | 0.34 | 0.78 |
| 0.68 | 0.74 | 0.65 | 0.49 | 0.73 | 0.77 | 0.80 |
| 0.76 | 0.60 | 0.80 | 0.42 | 0.79 | 0.41 | 0.84 |
| 0.78 | 0.68 | 0.79 | 0.43 | 0.78 | 0.42 | 0.75 |
| 0.83 | 0.83 | 0.74 | 0.33 | 0.82 | 0.73 | 0.93 |
| 0.74 | 0.87 | 0.83 | 0.50 | 0.78 | 0.43 | 0.84 |
| 0.85 | 0.57 | 0.82 | 0.76 | 0.72 | 0.47 | 0.79 |
| 0.68 | 0.49 | 0.68 | 0.33 | 0.77 | 0.77 | 0.73 |
| 0.68 | 0.74 | 0.76 | 0.43 | 0.82 | 0.38 | 0.92 |
| 0.74 | 0.57 | 0.77 | 0.70 | 0.67 | 0.41 | 0.79 |
| 0.65 | 0.57 | 0.70 | 0.35 | 0.73 | 0.38 | 0.69 |
| 0.68 | 0.82 | 0.69 | 0.40 | 0.62 | 0.37 | 0.77 |
| 0.49 | 0.50 | 0.48 | 0.49 | 0.48 | 0.33 | 0.48 |

| Pneumonitis Macro F1 | Myocarditis Binary F1 | Myocarditis Macro F1 | Dermatitis Binary F1 | Dermatitis Macro F1 | Cost/Note ($) |
|---|---|---|---|---|---|
| 0.67 | 0.86 | 0.67 | 0.92 | 0.73 | 0.02 |
| 0.59 | 0.77 | 0.70 | 0.90 | 0.68 | 0.00 |
| 0.50 | 0.73 | 0.46 | 0.77 | 0.42 | 0.00 |
| 0.49 | 0.84 | 0.34 | 0.79 | 0.68 | 0.00 |
| 0.54 | 0.74 | 0.70 | 0.85 | 0.62 | 0.00 |
| 0.61 | 1.00 | 0.67 | 0.78 | 0.56 | 0.02 |
| 0.33 | 0.73 | 0.33 | 0.81 | 0.57 | 0.00 |
| 0.63 | 1.00 | 0.67 | 0.86 | 0.56 | 0.01 |
| 0.47 | 0.76 | 0.54 | 0.78 | 0.38 | 0.02 |
| 0.48 | 0.77 | 0.39 | 0.83 | 0.66 | 0.00 |
| 0.60 | 0.49 | 0.83 | 0.89 | 0.50 | 0.01 |
| 0.46 | 0.79 | 0.23 | 0.73 | 0.64 | 0.00 |
| 0.52 | 0.82 | 0.68 | 0.68 | 0.53 | 0.00 |
| 0.58 | 0.74 | 0.50 | 1.00 | 0.50 | 0.02 |
| 0.51 | 0.70 | 0.40 | 0.73 | 0.66 | 0.00 |
| 0.50 | 0.70 | 0.53 | 0.64 | 0.60 | 0.00 |
| 0.37 | 0.70 | 0.32 | 0.81 | 0.71 | 0.00 |
| 0.47 | 0.75 | 0.59 | 0.73 | 0.56 | 0.00 |
| 0.32 | 0.48 | 0.33 | 0.49 | 0.33 | 0.00 |
