## Appendix Table 2 for "An agentic AI system enhances clinical detection of immunotherapy toxicities: a multi-phase validation study"

### Appendix Table 2: Pre/post survey results of AI-assisted labeling usability and attitudes

| Metric | Scale | Pre | Post | Δ | N | p-value |
|---|---|---|---|---|---|---|
| System Usability Scale (SUS) | 0–100 | 35.34 | 52.05 | 16.70 | 22 | **<0.001** |
| Perceived efficiency (interface) | 1–5 | 2.53 | 3.94 | 1.41 | 17 | **0.005** |
| Perceived label accuracy confidence (interface) | 1–5 | 3.12 | 3.82 | 0.71 | 17 | **0.018** |
| Confidence AI can assist (task) | 1–5 | 2.27 | 3.00 | 0.65 | 17 | **0.026** |
| Preference: AI vs Manual (post) | % prefer AI | — | 88.20 | 82.40 | 17 | — |

*SUS row uses a paired t-test on total SUS scores (0–100). Likert-scale items use paired Wilcoxon signed-rank tests after ordinal mapping (1–5). The preference row reports the percent of participants preferring AI and the margin vs. manual; it is post-only and no paired test is applicable. Abbreviations: SUS = System Usability Scale; Δ = mean change (Post–Pre).*
