## Appendix 1 for "An agentic AI system enhances clinical detection of immunotherapy toxicities: a multi-phase validation study"

**Participant Demographics and Characteristics for those who completed the HCI Study**

| | | |
|---|---|---|
| **Total Participants** | 17 | |
| **Primary Role** | | |
| Clinical research coordinator | 16 (94.1%) | |
| Nurse | 1 (5.9%) | |
| **Years in Oncology Research** | | |
| Mean (SD) | | 2.3 (1.9) |
| Median (Range) | | 2.0 (0.5-8.0) |
| **CTCAE Grading Experience** | | |
| Slight | 5 (31.2%) | |
| Moderate | 5 (31.2%) | |
| Extensive | 4 (25.0%) | |
| Expert | 2 (12.5%) | |
| **irAE Identification Experience** | | |
| Slight | 3 (27.3%) | |
| Moderate | 6 (54.5%) | |
| Extensive | 1 (9.1%) | |
| Expert | 1 (9.1%) | |
| **Prior AI Experience (General)** | | |
| Slight | 6 (42.9%) | |
| Moderate | 7 (50.0%) | |
| Extreme | 1 (7.1%) | |
| **Trust in LLMs for Clinical Information** | | |
| Slight | 11 (73.3%) | |
| Moderate | 2 (13.3%) | |
| Very | 1 (6.7%) | |
| Extreme | 1 (6.7%) | |
| **LLM Use for Work-Related Tasks** | | |
| Never | 7 (41.2%) | |
| Rarely | 4 (23.5%) | |
| Sometimes | 3 (17.6%) | |
| Often | 1 (5.9%) | |
| Very Often / Routinely | 2 (11.8%) | |
| **Belief AI Can Benefit Medicine (Long-term)** | | |
| Neutral | 3 (17.6%) | |
| Agree | 12 (70.6%) | |
| Strongly Agree | 2 (11.8%) | |
