## Appendix 2 for "An agentic AI system enhances clinical detection of immunotherapy toxicities: a multi-phase validation study"

**Appendix 2. Patient-level demographic information across the 3 validation stages**

| Characteristic | | Field cohort (N=27) | Prospective cohort (N=228) | Retrospective cohort (N=100) |
|---|---|---|---|---|
| Age (years) | | 67.4 (8.7) | 63.9 (14.6) | 66.3 (13.2) |
| Sex | Female | 17 (63.0%) | 114 (50.0%) | 55 (55.0%) |
| | Male | 10 (37.0%) | 114 (50.0%) | 45 (45.0%) |
| Race group | White | 27 (100.0%) | 195 (85.5%) | 97 (97.0%) |
| | Black | 0 (0.0%) | 14 (6.1%) | 1 (1.0%) |
| | Asian | 0 (0.0%) | 5 (2.2%) | 2 (2.0%) |
| | Other | 0 (0.0%) | 7 (3.1%) | 0 (0.0%) |
| | Declined | 0 (0.0%) | 4 (1.8%) | 0 (0.0%) |
| | Two or More | 0 (0.0%) | 1 (0.4%) | 0 (0.0%) |
| Ethnic group | Non Hispanic | 27 (100.0%) | 213 (93.4%) | 100 (100.0%) |
| | Declined | 0 (0.0%) | 7 (3.1%) | 0 (0.0%) |
| | Hispanic | 0 (0.0%) | 4 (1.8%) | 0 (0.0%) |
| Primary language | English | 27 (100.0%) | 208 (91.2%) | 96 (96.0%) |
| | Non-English | 0 (0.0%) | 20 (8.8%) | 4 (4.0%) |

Age is presented as the mean (SD), with the rest shown as exact percentages rounded to 1dp. Note, multiple notes may be present per patient.
