## Appendix 3 for "An agentic AI system enhances clinical detection of immunotherapy toxicities: a multi-phase validation study"

### Immune-Related Adverse Event Annotation Guidelines


1. Introduction and Purpose Statement 1
2. Annotation Task 2
3. Temporality 5
4. irAEs of interest 7
i. Myocarditis 7
ii. Dermatitis 8
iii. Colitis 8
iv. Hepatitis 8
v. Thyroiditis 9
vi. Pneumonitis 10
5. Attributes 11
a. Severity 11
b. Attribution 12
c. Certainty 12
6. Notes and Edge Cases 14
d. Certainty 14
e. Contradictory Information 14
f. Negated entities 14
g. Copy-pasted radiology reports 14
h. Laboratory values 14
i. Imaging findings 15
j. Hypothyroidism 15
Appendix I: CTCAE term mappings 16

### 1. Introduction and Purpose Statement

Our ability to understand cancer treatment-related adverse effects, including immune-related adverse events, is limited by a lack of data. Cancer treatment side effects are not collected in most cancer registries, and clinical trials are inherently limited in their ability to collect adverse event data. The goal of this project is to develop natural language processing (NLP) methods that can automatically extract information about irAEs in adult cancer patients. The ultimate goal is to use these methods to create real world evidence on irAEs, and to enable technologies to automatically monitor patients' electronic health records (EHRs) for these events.

These guideline describe specific irAEs we wish to capture in the clinical documents of patients undergoing or completed immunotherapy, and the methodology for annotation. In addition to identifying mentions of disorders that may be irAEs in documents, each irAE will have certain attributes to further enrich our datasets.

The annotations developed in this project will subsequently be used to train NLP systems that can "read" clinical documents, identify toxicities of interest, and extract them from the texts. NO EXAMPLES IN THIS DOCUMENT ARE PHI – THEY ARE NOT REAL CLINICAL TEXT.

**Annotators should carefully read these guidelines in their entirety prior to starting annotations.** Please adhere as closely to these guidelines, even in cases that you disagree with definitions or rules provided. This is important to ensure annotation quality and consistency.

### 2. Annotation Task

The annotation task to provide each document with labels that provide information about the presence of irAEs, along with descriptive attributes. *Do not use information from other notes on the same patient to arrive at a label.*

Please note that the irAEs and their attributes may not be present in the note as described verbatim below. There are varied natural language expressions of each class, so please use best judgement in determining if the class or attribute is present.

We will start labeling only the 6 priority disorders for now: **Myocarditis, dermatitis, thyroiditis, hepatitis, colitis, and pneumonitis.** It is important to note that these irAEs are umbrella terms for multiple CTCAE terms. For example, immune-related cough should be labeled as pneumonitis. The term mappings are in Appendix I: CTCAE term mappings. All notes were selected such that the note was written within 12 months of an immunotherapy infusion. You may use this information to make a determination about the irAE.

Important details to guide the labeling are as follows:

1. The goal is to label medical events for which the irAE is in the differential diagnosis, not every single isolated sign or symptom that may overlap with an irAE. These irAEs are generally defined by clusters of signs/symptoms that, taken together, are

suggestive of the disorder. Often, there will be other diagnoses in the differential at the time the note is written. As long as the irAE is in the differential, you should still label the irAE if it is an unlikely or possible diagnosis, unless there is a clear alternative diagnosis (see 2 below). I have added lists of signs and symptoms that are often seen with of each of the 6 priority irAEs to the annotation guidelines, as well as common diagnostic procedures – these can guide whither an irAE is in the differential, even if the exact term is not used in the note

2. If the presentation is similar to an irAE but there is *clearly another diagnosis that explains the presentation, **and** it is stated as such in the note, do not label the irAE is present.* For example, if a patient is already diagnosed with an infectious pneumonia and the note says that the pneumonia is the cause of shortness of breath or fever, no need to label dyspnea and fever or any associated context spans – the dyspnea and fever are not describing a pneumonitis event. If a patient has chronic shortness of breath that is clearly attributed to longstanding COPD, no need to label it – the shortness of breath is not describing a pneumonitis event. If a patient is having chest pain but it is clearly because of coronary artery disease, no need to label it – the chest pain is not describing a myocarditis event.

The below table describes the labels and their value sets for this project. For each irAE type and temporality (current vs. past) combination, we label its presence/absence, CTCAE grade, attribution, and certainty. Temporality and irAE types are described in more detail in Sections 3 and 4, respectively. The Attributes section provides details for the irAE type, Grade, Attribution, and Certainty labels. Please read through these details carefully and reference them as you annotate. They include detailed examples and instructions to guide your annotations.

| **Label** | **Name** | **Description** | **ValueSet** |
|---|---|---|---|
| CurrentType_* | Current irAE Type | At the time the note is written, is the patient currently experiencing the irAE? Label yes if the irAE is in the differential, even if it is not certain. Details: irAEs of Interest | 1=yes; blank = no |
| CurrentGrade_ | Current **Grade** | If yes to CurrentType, what is the current CTCAE grade of that irAE? Details: Severity | 1=grade 1; 2=grade 2; 3=grade 3; 4=grade 4; 5=grade 5 |
| CurrentAttr_ | Current **Attribution** | If yes to CurrentType, what is the Attribution? Details: Attribution | 1 = IO; blank = not IO |
| CurrentCert_ | Current **Certainty** | If yes to CurrentType & CurrentAttr, what is the explicit certainty that the event is due to immunotherapy? Details: Certainty | 1 = unlikely; 2 = Possible; 3 = Likely; 4 = Certain; blank = no explicit certainty described |
| PastType_ | Past irAE Type | Did the patient have that irAE type in the past? Label yes if the irAE was in the differential and no other clear alternative diagnosis was determined, even if it was not certain. Details: irAEs of interest | 1=yes; blank = no |
| PastMaxGrade_ | Past Maximum **Grade** | If yes to PastType, what was the maximum grade (not counting the current grade, if relevant)? Details: Severity | 1=grade 1; 2=grade 2; 3=grade 3; 4=grade 4; 5=grade 5 |
| PastAttr_ | Past **Attribution** | If yes to PastType, what is the Attribution? Details: Attribution | 1 = IO; blank = not IO |
| PastCert_ | Past **Certainty** | If yes to PastType & PastCert; what is the explicit certainty that the event is due to immunotherapy? Details: Certainty | 1 = unlikely; 2 = Possible; 3 = Likely; 4 = Certain; blank = no explicit certainty described |

*Each label is appended by an abbreviation for the irAE type in question: Pneum, Col, Derm, Hep, Thyr, Myo

### 3. Temporality

There are different ways irAEs may be described over time which largely break down into the following buckets:

1. Past, now resolved irAEs: An irAE that started in the past and is now completely resolved. This includes irAEs that recurred multiple times in the past, but are now resolved. These events should receive only Past* labels.
    a. Note thate we usually hold immunotherapy permanently after someone experiences and irAE, even if the irAE resolves. Therefore, text such as “stage I lung adenocarcinoma (previously on pembro but on hold 2/2 pneumonitis)” on its own is insufficient to label pneumonitis as Currently present. It should only get Past labels.
2. Ongoing irAEs: An irAE that started in the past, and is still ongoing. The severity may have changed over time, it may have also resolved and recurred multiple times in the past, and/or it may be a chronic condition (usually the case for endocrinopathies). These events should receive both Current* and Past* labels.
    a. If patient still on immune suppressants for the irAE but not symptomatic, the irAE is still present so label it as Current.
    b. If a patient was admitted to one hospital with the irAE and transferred to the current hospital, then it is both Past and Current because it is not the firs time the patient has presented for medical care due to that condition (even if it is the first time presenting to the current institution)
3. New irAE presentations: The patient is presenting for the first time ever with the irAE. These events should only receive Current* labels.

Detailed timeline extraction is out-of-scope this project. For the Current_* labels, label what is occurring to that patient *right now* (at the time the note is written). For example, if a patient had a past grade 3 irAE, which is now better but still on steroids, the CurrentGrade = 2 and PastGrade =3. If a patient had an irAE that is now resolved, it does not get the Current* labels.

For the PastGrade, label the past maximum grade experienced, *not counting the current grade*. For example if a patient had a past maximum grade 2 irAE, and now is admitted with a grade 3 irAE, CurrentGrade = 3 and PastGrade = 2.

Sometimes, the language is ambiguous as to whether the irAE is past or present. A common description that is ambiguous is language such as: “The patient is s/p immunotherapy, course complicated by pneumonitis”. Unless it is otherwise clear elsewhere in the note that the irAE is ongoing, this should labeled as a Past event only.

One edge area is when the patient experienced multiple episodes of the same irAE type over time, but they are described with differing Certainty and Attributes. I find it is usually quite easy to resolve these issues. Here, use your best common sense – if one of the events is described with a clear Certainty and Attribution, and then a new event is clearly and explicitly described as a recurrence of the prior event without but less details are provided, then favor giving the Certainty and Attribute labels from the prior event it is being to linked to.

- Examples
  - "The patient previously had immunotherapy-related colitis treated with steroids. The patient now presents with recurrence of the colitis." *The current event would be labeled with IO = yes and Certainty = certain because it is clearly described as a recurrence of the past event where clearly IO = yes and Certainty = certain:*
    - CurrentType_col = yes
    - CurrentGrade_col=1 (max inferrable grade based on this text)
    - CurrentAttr_col = yes
    - CurrentCert_col = yes
    - PastType_col = yes
    - PastGrade_col = 2
    - PastAttr_col = yes
    - PastCert_col = yes
  - "The patient previously had immunotherapy-related colitis treated with steroids. The patient now presents with possible recurrence of colitis." *The word recurrence makes it clear that the current colitis described in orange refers to the past colitis referred to in purple. The past colitis (purple) is Attribution = yes, and so the current colitis (orange) inherits this same attribution, However, the Certainty is different for the past and present event.*
    - CurrentType_col = yes
    - CurrentGrade_col=1 (max inferrable grade based on this text)
    - CurrentAttr_col = yes
    - CurrentCert_col = no
    - PastType_col = yes
    - PastGrade_col = 2
    - PastAttr_col = yes
    - PastCert_col = yes
  - "Previously admitted for pembro-related pneumonitis… now is admitted for dyspnea due to pneumonia vs inflammatory process such as pneumonitis (2/2 ?too quick discontinuation of steroid taper for prior pneumonitis.)"
    - CurrentType_Pneum = 1
    - CurrentGrafe_Pneum = 3 (because admitted to the hospital for this event)
    - CurrentAttr_Pneum = 1
    - CurrentCert = 2 (possible) because the prior pneumonitis is attributed certainly to pembrolizumab, but this event is only possibly a recurrence of the pembro-induced pneumonitis (because of the "vs"). We know that the

possible pneumonitis is felt to be a recurrence of the prior pembro-related pneumonitis because of “2/2 ?too quick discontinuation of steroid taper for prior pneumonitis”

- PastType_Pneum = 1
- PastGrade_Pneum = 3
- PastAttr_Pneum = 1
- PastCert_Pneum = 4

### 4. irAEs of interest

These are the types of irAEs that we are annotating. Descriptions of the irAEs below are adapted from the A151804 protocol. We are labeling the following irAEs: myocarditis, pneumonitis, dermatitis, hepatitis, colitis, thyroiditis. We may extend to other irAEs in the future (descriptions of out-of-scope irAEs are in Appendix II). It is important to note that these irAEs are umbrella terms for multiple CTCAE terms. For example, immune-related cough should be labeled as pneumonitis. The term mappings are in Appendix I: CTCAE term mappings.

Many signs, symptoms, diseases, and disorders commonly seen for the irAEs of interest are described in Section 5 above. Please refer to the Alliance A151804 irAE to CTCAE v5 term mappings in Appendix I, which provide additional terms which may refer to or are subsumed within the below irAEs. Other resources to help identify adverse event anchors include: https://jitc.bmj.com/content/9/6/e002435.

#### i. Myocarditis

The diagnosis of myocarditis is challenging since the clinical manifestations are nonspecific. Signs and symptoms include chest pain, dyspnea and arrhythmia. Diagnostic tests used to diagnose myocarditis include troponin (a lab), electrocardiograms, cardiac MRIs, and biopsies. Troponin elevation is common but is nonspecific. Electrocardiograms may show subtle, non-specific changes or be normal. Cardiac MRI with contrast is more sensitive. Endomyocardial biopsy is considered the gold standard.

Below is a list of signs and symptoms suggestive of myocarditis or its sequelae and should be labeled as anchors unless there is another clear diagnosis that explains them (please also refer to the terms in Appendix I: CTCAE term mappings:

Cardiovascular signs/symptoms:

- Chest pain or discomfort (often but not always sharp, pressure-like, worse with breathing or lying down)
- Shortness of breath (dyspnea)
- Palpitations (irregular or rapid heartbeat)/arrhythmia (see lists of arrythmia types in Appendix I: CTCAE term mappings)

- Fainting (syncope): Often due to irregular or rapid heartbeat
- Dizziness/lightheadedness/pre-syncope: Often due to irregular or rapid heartbeat
- Elevated troponin
- EKG QT corrected interval prolonged
- Conduction disorder (see specific conduction disorders in Appendix I: CTCAE term mappings)
- Cardiac MRI abnormality

Symptoms associated with heart failure, which may occur secondary to myocarditis

- Swelling (edema)/fluid retention (often manifests as lower extremity swelling, weight gain)
- Orthopnea (shortness of breath when lying flat)
- Paroxysmal nocturnal dyspnea (sudden shortness of breath during sleep)

#### ii. Dermatitis

Signs and symptoms of dermatitis include rash and pruritus. Biopsy can be performed unless contraindicated.

Below is a list of signs/symptoms suggestive of dermatitis and should be labeled as anchors unless there is another clear diagnosis that explains them (please also refer to the terms in Appendix I: CTCAE term mappings:

- Rash
- Skin redness (erythema)
- Itching (pruritis)
- Dry skin (including flaking/cracked skin)
- Swelling (edema) in the affected area
- Pain, tenderness, or sensitivity in the affected area
- Scaling or peeling skin

#### iii. Colitis

Signs and symptoms of colitis include diarrhea, abdominal pain, mucus or blood in stool, with or without fever, and those of bowel perforation including peritoneal signs and ileus. Infectious etiologies such as Clostridium difficile is often part of the differential diagnosis. Abdominal CT may be helpful to diagnose colitis. Sigmoidoscopy or colonoscopy with biopsy should be considered for persistent or severe symptoms.

Below is a list of signs/symptoms suggestive of colitis and should be labeled as anchors unless there is another clear diagnosis that explains them (please also refer to the terms in Appendix I: CTCAE term mappings:

- Diarrhea, frequent, loose, or watery stools (often a hallmark)
- Abdominal pain or cramping
- Bloody stools
- Mucus in stools
- Peritonitis
- Loss of appetite
- Ileus symptoms: Constipation, inability to pass gas, hypoactive bowel sounds, nausea/vomiting

#### iv. Hepatitis

Symptoms of hepatitis include right upper-quadrant abdominal pain and/or unexplained nausea or vomiting. Laboratory abnormalities indicative of immune-mediated hepatitis include elevated hepatic transaminases and bilirubin levels. Infectious or malignant are frequent differential diagnoses.

Below are lists of signs/symptoms suggestive of hepatitis and should be labeled as anchors unless there is another clear diagnosis that explains them (please also refer to the terms in Appendix I: CTCAE term mappings.

Acute inflammatory hepatitis:

- Nausea/vomiting
- Abdominal pain (especially in the right upper quadrant)
- Bloating or fullness
- Hepatomegaly (enlarged liver, palpable on exam or seen on imaging)
- Elevated transaminases
- Elevated bilirubin

Advanced disease/liver failure:

- Ascites (fluid buildup in abdomen)
- Peripheral edema
- Easy bruising and bleeding

- Confusion or mental changes (encephalopathy)
- Jaundice (yellowing of skin or eyes)
- Dark urine
- Pale or clay-colored stools
- Pruritis (itching)
- Muscle wasting

#### v. Thyroiditis

Immune-related thyroiditis can present as hyper- or hypothyroidism. TSH and free T4 levels are used to determine whether thyroid abnormalities are present. Thyroiditis patients may be hyperthyroid, euthyroid, hypothyroid and typically progress through these phases. Typically, hypothyroid is the final and chronic state after thyroiditis has "burnt-out".

**Importantly and uniquely,** because hypothyroidism is common in the general population, only label if there is some reason to believe per the note that immune-related hypothyroidism is in the differential diagnosis for hypothyroidism.

Below are lists of signs/symptoms suggestive of thyroiditis and should be labeled as anchors unless there is another clear diagnosis that explains them (please also refer to the terms in Appendix I: CTCAE term mappings:

General symptoms during the acute active thyroiditis phase (patients are generally hyperthyroid during this phase):

- Neck pain or tenderness (localized over thyroid gland, may radiate to jaw/ears)
- Neck swelling (goiter): Visible or palpable thyroid enlargement

Hyperthyroid signs/symptoms:

- Low TSH, elevated free T4
- Rapid Heartbeat (Tachycardia)
- Nervousness or Anxiety
- Heat Intolerance
- Increased Sweating
- Unexplained weight loss
- Tremors
- Increased Appetite

- Insomnia
- Frequent Bowel Movements or Diarrhea
- Irritability or Restlessness
- Menstrual Irregularities
- Hair Thinning

Hypothyroid signs/symptoms:

- Elevated TSH, low free T4
- Cold Intolerance
- Unexplained weight gain
- Dry Skin
- Hair Loss
- Constipation
- Slow Heart Rate (Bradycardia)
- Depression
- Memory Problems or Brain Fog
- Hoarseness
- Puffy Face or Swelling and thickening of skin (Myxedema): A severe and uncommon form of hypothyroidism
- Muscle Weakness or Cramps
- Joint Pain or Stiffness
- Prolonged Menstrual Periods (Menorrhagia)

#### vi. Pneumonitis

Signs and symptoms of pneumonitis include dyspnea, cough, fatigue and hypoxia. Onset varies widely with a median of 2.5 months (range, 0.8–11 months). Other etiologies that are often in the differential include pneumonia, lymphangitic carcinomatosis, pulmonary embolism, heart failure, COPD or pulmonary hypertension. Common assessments include measurement of oxygen saturation, high-resolution chest CT, pulmonary function tests including spirometry and DLCO, and bronchoscopy with bronchoalveolar lavage and biopsy.

Below is a list of signs/symptoms suggestive of thyroiditis and should be labeled as anchors unless there is another clear diagnosis that explains them (please also refer to the terms in Appendix I: CTCAE term mappings:

Common signs/symptoms:

- Cough (often a dry cough)
- Shortness of breath (dyspnea)
- Hypoxia
- Chest pain with deep breaths (pleuritic)
- Wheezing
- Cyanosis
- Tachypnea/increased respiratory rate

### 5. Attributes

#### a. Severity

Label the *maximum severity* that can be inferred from information in the document. In cases of ambiguity, lean toward selecting the higher severity grade. Severity grading is different for each irAE (Grade 5 always refers to death). Please refer to the CTCAE v5 to determine severity, on a grade of 1-5: https://ctep.cancer.gov/protocoldevelopment/electronic_applications/docs/ctcae_v5_quick_reference_5x7.pdf. Importantly, not all irAE terms above appear in the CTCAE guidelines. **Appendix I: CTCAE term mappings** provides mappings for each irAE to the relevant CTCAE terms.

A cheatsheet to the CTCAE guidelines for our target irAEs is here: https://docs.google.com/document/d/19jirSz98MPTaW2HBux6RYyQNhyMngs2eUdpG5_NvJBQ/edit?usp=sharing

Always remember to give a grade if the irAE is present. Can default to Grade = 1 if there is otherwise not enough info to give a higher grade. For example, if all we know about the irAE is that it caused the IO to be discontinued or put on hold, Grade = 1.

Note: Cannot call grade >1 based on imaging findings alone, even if imaging findings are extensive. However, mention of symptoms/medications/hospitalizations in the order or the impression can be taken into account when determining grade.

- **Grade 1:** Refer to CTCAE guidelines
- **Grade 2:** Refer to CTCAE guidelines
    - Note that oral steroids usually mean Grade 2, however topical steroids alone do not mean Grade 2 (e.g., someone can be on topical steroids for rash and have grade 1 dermatitis)

- if there is mention that the patient has symptoms related to imaging finding in the radiology report, it can be considered grade 2

- **Grade 3:** Refer to CTCAE guidelines
    - If someone is admitted to the hospital due to the irAE, then Grade = 3.
    - If someone is admitted to the hospital for something else but also has an irAE, and the irAE is delaying discharge from hospital, Grade = 3
- **Grade 4:** Refer to CTCAE guidelines
- **Grade 5:** Death

#### b. Attribution

The Attribution property indicates whether the irAE is explicitly attributed to immunotherapy by the note author. Determine Attribution based only on explicit documentation by the note writer that the irAE is caused by an immunotherapy or not, e.g., "The patient's pneumonitis is likely due to immunotherapy". **Mentions of "drug-related" only counts as IO if it is clear that the "drug" refers to the IO – note that many drugs can cause inflammatory adverse events. Please note that if is explicitly stated that an IO was discontinued due to the event, then that counts as attribution = IO because this means the clinician linked this event to the IO. Note that Not IO is the default label, only add a label if attribution is IO.** Attribution labels are as follows:

- **IO:** The Disorder is explicitly described as being caused by immunotherapy by the note author.
    - Examples:
        - "The patient had immune-related nephritis"
        - "Patient recently started on nivolumab… presenting with drug-related colitis due to his recent IO treatment".
        - "Pembro was discontinued due to pneumonitis"

- **Not IO:** The disorder is not explicitly attributed to IO. This is the default label.
    - Example:
        - "Patient had a rash after 2nd infusion of nivolumab" - This is None even though the timing seems like it should be due to immunotherapy, because the clinician does not explicitly say that the rash is *caused by* immunotherapy

#### c. Certainty

Certainty refers to *explicit* mentions of the clinician's certainty that the event is related to immunotherapy. It does not refer to the annotator's certainty. Terms here mirror the terminology used in the Naranjo scale (https://www.ncbi.nlm.nih.gov/books/NBK548069/). **Note that None is the default label, only add a label if there is an explicit certainty level described.**

- **None:** The event is explicitly attributed to immunotherapy, but there is described no level of certainty that the event is related to immunotherapy [DEFAULT]
    - "Pembro was discontinued due to pneumonitis"
        - While this suggests the provider thought the event was due to immunotherapy, the certainty is not clear as they may have discontinued even though there was uncertainty.
- **Unlikely (Doubtful):** Includes situations where the clinician believes that it is unlikely or doubtful to be immunotherapy-related.
    - Examples:
        - "There is a chance that the symptoms are related to immunotherapy but it is more likely to be a COPD exacerbation."
        - "Immune-related dermatitis is in the differential but unlikely given the timing."
        - "The imaging findings on chest CT might be related to nivolumab, but looks more consistent with radiation-related changes."

- **Possible:** Descriptions of the event as possibly related to immunotherapy. This includes situations where it is expressed that the different etiologies are equally likely. "Concerning for" = possible. "Suspected" = possible.
    - Examples:
        - "This could be related to radiotherapy or immunotherapy."
        - "Symptoms are concerning for immune-related colitis."
        - "Ddx ICI colitis vs infection"

- **Probably (Likely):** This includes situations where immunotherapy is described as the likely cause, the most likely cause, or the leading cause. This includes when the clinician "feels" it is the cause of the event or "thinks"/"presumes" it is the cause of the event.
    - Examples:
        - **"**The TSH elevation is likely caused by ipi/nivo."
        - "This is in all likelihood immune-related EKG changes."
        - "Immune-related transaminitis elevation is the leading diagnosis."
        - "This is most consistent with immune-related pneumonitis" *Note that "most consistent with" is Probably, while "consistent with" without qualifiers is Certain.*
        - "Pulmonology feels this is pembro-induced pneumonitis" or "Pulmonology thinks this is pembro-induced pneumonitis"
        - "Recent admissions for colitis thought to be 2/2 CPI"

    - “Admitted from clinic for presumed ICI myocarditis”

- **Certain (Definite):** The clinician is absolutely clear/certain/definite that the event is related to immunotherapy. Alternatively, the event is clearly and explicitly diagnosed as an immune-releted event. If a provider describes treating the condition with an immunosuppressant but does not clearly express that it is immune-related, it should be labeled “None” because the certainty is not explicit.
  - Examples:
    - “This is immune-related pneumonitis.”
    - “He was on nivolumab complicated by grade III skin toxicitiy”
    - “This is consistent with immune-related pneumonitis”

- **None:** The clinician’s certainty is not explicitly mentioned. This is the default label.
  - Examples:
    - **“**I will prescribe steroids for pneumonitis.”

### 6. Notes and Edge Cases

#### d. Certainty

Statements of certainty (or lack thereof) may complicate labeling and may require the annotator’s best judgement. Please use your best judgement. Also refer to Certainty, which is an explicit label for the clinician’s certainty that an event is caused by immunotherapy.

#### e. Contradictory Information

It is possible a document may contradict itself. In these cases, please use your best judgement but in general preference the positive/present/more severe labels.

#### f. Negated entities

Do not label description of a disorder not being present. For example, “The patient has no pneumonitis.” should not be labeled.

#### g. Copy-pasted radiology reports

Radiology reports or sections of reports are often copy-pasted into other clinical notes. In some cases, these describe an irAE. If the label(s) based on the radiology report disagrees with the clinical note label(s), preference the clinical note label(s). For CTCAE Grade 1 events, there may be only mention of the irAE being present or possible present in the radiology report. In these cases, you should label according the radiology report even if the irAE is not mentioned elsewhere in the note.

##### h. Laboratory values

Do not use lab values in isolation to label irAEs.

##### i. Imaging findings

Explicit descriptions that the imaging findings demonstrate an inflammatory reaction or immune-mediated reaction can be considered an irAE (e.g., “immune-mediated process in the lung” would be labeled pneumonitis). However, “ground-glass opacities”, “stranding”, or “consolidations”, without additional descriptions of these indicate, are insufficient to be considered the irAE.

##### j. Hypothyroidism

Importantly and uniquely, because hypothyroidism is common in the general population, only label if there is some reason to believe per the note that immune-related hypothyroidism is in the differential diagnosis for hypothyroidism.

### Appendix I: CTCAE term mappings

Alliance A151804 irAEs of interest, and related AEs listed in CTCAE v5, include:

| **irAE** | **CTCAE MedDRA Code** | **CTCAE v5 terms** |
|---|---|---|
| Myocarditis | 10003586<br>10003658<br>10003662<br>10003673<br>10007515<br>10007541<br>10010276<br>10019279<br>10069501<br>10027786<br>10027787<br>10028596<br>10028606<br>10034040<br>10034474<br>10053565<br>10034484<br>10058597<br>10040639<br>10042604<br>10047281<br>10047290<br>10047302<br>10007612<br>10007613<br>10014383 | Asystole<br>Atrial fibrillation<br>Atrial flutter<br>Atrioventricular block complete<br>Cardiac arrest<br>Cardiac disorders – Other, specify<br>Conduction disorder<br>Heart failure<br>Left ventricular systolic dysfunction<br>Mobitz (type) II atrioventricular block<br>Mobitz type I<br>Myocardial infarction<br>Myocarditis<br>Paroxysmal atrial tachycardia<br>Pericardial effusion<br>Pericardial tamponade<br>Pericarditis<br>Right ventricular dysfunction<br>Sick sinus syndrome<br>Supraventricular tachycardia<br>Ventricular arrhythmia<br>Ventricular fibrillation<br>Ventricular tachycardia<br>Cardiac troponin I increased<br>Cardiac troponin T increased<br>Electrocardiogram QT corrected interval prolonged |
| Colitis | 10000081<br>10065747<br>10009887<br>10009998<br>10012727<br>10014893<br>10021328<br>10038064 | Abdominal pain<br>Cecal hemorrhage<br>Colitis<br>Colonic hemorrhage<br>Diarrhea<br>Enterocolitis<br>Ileus<br>Rectal hemorrhage |
| Hepatitis | 10001551<br>10001675<br>10003481<br>10005364 | Alanine aminotransferase increased<br>Alkaline phosphatase increased<br>Aspartate aminotransferase increased<br>Blood bilirubin increased |
| Nephritis | 10069339 | Acute kidney injury |

| | | |
|---|---|---|
| | 10011368 | Creatinine increased |
| | 10019450 | Hematuria |
| | 10037032 | Proteinuria |
| Myositis | 10011268 | CPK increased |
| | 10062572 | Generalized muscle weakness |
| | 10065776 | Muscle weakness lower extremity |
| | 10065795 | Muscle weakness trunk |
| | 10065895 | Muscle weakness upper extremity |
| | 10028411 | Myalgia |
| | 10028653 | Myositis |
| Pneumonitis | 10011224 | Cough |
| | 10013963 | Dyspnea |
| | 10021143 | Hypoxia |
| | 10035742 | Pneumonitis |
| Meningitis/Encephalitis | 10014625 | Encephalopathy |
| | 10019211 | Headache |
| | 10027198 | Meningismus |
| Dermatitis | 10006556 | Bullous dermatitis |
| | 10015218 | Erythema multiforme |
| | 10015277 | Erythroderma |
| | 10037868 | Rash maculo-papular |
| | 10042033 | Stevens-Johnson syndrome |
| | 10044223 | Toxic epidermal necrolysis |
| Endocrinopathies | 10001367 | Adrenal insufficiency |
| | 10020639 | Hyperglycemia |
| | 10020850 | Hyperthyroidism |
| | 10062767 | Hypophysitis |
| | 10021114 | Hypothyroidism |
| | 10043770 | Thyroid stimulating hormone increased |
| Neuropathy | 10018767 | Guillain-Barre syndrome |
| | 10065780 | Muscle weakness left-sided |
| | 10065794 | Muscle weakness right-sided |
| | 10033987 | Paresthesia |
| | 10034580 | Peripheral motor neuropathy |
| | 10034620 | Peripheral sensory neuropathy |
| Other rheumatological | 10003239 | Abducens nerve disorder |
| | 10003246 | Facial muscle weakness |
| | 10023215 | Facial nerve disorder |
| | 10048706 | Arthralgia |
| | 10065796 | Arthritis |
| | 10065800 | Joint effusion |
| | 10028395 | Joint range of motion decreased |
| | 10033425 | Joint range of motion decreased cervical spine |
| | 10053662 | Joint range of motion decreased lumbar spine |
| | 10051272 | Musculoskeletal and connective tissue disorder - Other, specify |
| | 10061457 | Myasthenia gravis |
| | 10028417 | Pain in extremity |
| | 10047115 | Vasculitis |

| | | |
|---|---|---|
| Hematologic cytopenias | 10002272<br>10016288<br>10025256<br>10029366<br>10035528<br>10049182 | Anemia<br>Febrile neutropenia<br>Lymphocyte count decreased<br>Neutrophil count decreased<br>Platelet count decreased<br>White blood cell count decreased |
| Pancreatitis | 10000081<br>10028813<br>10033645<br>10047700<br>10024574<br>10040139<br>10003988 | Abdominal pain<br>Back pain<br>Lipase increased<br>Nausea<br>Pancreatitis<br>Serum amylase increased<br>Vomiting |
| Rare infections | | N/A |
| Hyperprogression | | N/A |
| Other | | Autoimmune disorder |
