## Appendix 4 for "An agentic AI system enhances clinical detection of immunotherapy toxicities: a multi-phase validation study"

### Appendix 4: Preparatory Field Study for Interface Design and Study Planning

#### Purpose

Prior to the randomized user effect study (Phase 3), we conducted a preparatory field study to (1) refine the annotation interface through iterative user feedback, (2) establish baseline efficiency and agreement metrics for power calculations, and (3) assess the temporal stability of human-AI collaboration over extended use.

#### Methods

##### *Interface Development*

We developed a web application integrating the agentic LLM system into annotation workflows, built using React 18.2 with a FastAPI backend. We conducted think-aloud sessions with 3 clinical research coordinators and iteratively incorporated their feedback to optimize interface design. Key refinements included evidence highlighting within the clinical note viewer, keyboard shortcuts for efficient navigation, and pre-population of structured data entry forms with model suggestions. Audit trails tracked human interactions, timings, and modifications.

##### *Field Deployment*

Two clinical research coordinators used the application over 3 months to assist chart curation for a separate retrospective study. We tracked annotation time per note, inter-annotator agreement, and human-model agreement over time. Results were reported descriptively.

#### Results

A total of 299 notes were co-annotated over six sequential time periods (approximately weekly buckets). Inter-rater agreement on grade was substantial overall (Cohen's $\kappa = 0.80$) and largely steady across time, with a transient dip for notes 101–150 ($\kappa = 0.55$) that rebounded thereafter ($\kappa = 0.71$–$0.93$). Agreement with the model on grade was high overall (mean $\kappa = 0.89$) and stable across time periods.

Time per note averaged 161 seconds versus an estimated 600-second baseline without AI assistance (~3.7× faster). Efficiency improved over time, demonstrating a learning curve: annotation times decreased from the first to final time periods for both annotators, and the between-annotator variability narrowed substantially.

#### Implications for Randomized Study Design

These findings informed several aspects of the Phase 3 randomized study:

1. Primary endpoint: Annotation time showed sufficient variability and meaningful differences to serve as the primary efficiency endpoint
2. Sample size: Observed effect sizes and variance estimates informed power calculations
3. Interface features: Evidence highlighting and keyboard navigation were retained based on positive user feedback
4. Training protocol: A one-hour training session was established as sufficient based on observed learning curves
