## Appendix 5a for "An agentic AI system enhances clinical detection of immunotherapy toxicities: a multi-phase validation study"

### Training Materials: Immune-Related Adverse Event (irAE) Labeling Study

#### Evaluating Language Model Assistance in Clinical Note Annotation

**Welcome!**

These are the training materials for participants in our human-computer interaction study. This study evaluates the impact of using a language model-aided labeling tool compared to our standard control method (reviewing text and using Google Sheets) for annotating immune-related adverse events (irAEs) in clinical notes.

This document provides essential information about the study, the annotation task, and the interfaces you will use. Please read it thoroughly before the training session. You will have the opportunity to ask questions during the session. This document can also be referenced throughout your participation.

**This document is organized into three main parts:**

1. **Study Information:** Details about the study structure and logistics.
2. **Annotation Guidelines:** Instructions on identifying and labeling irAEs and their associated attributes.
3. **Labeling Interface Overview:** Brief description of the tools you will use.
    a. https://irae-app-cqf0czf3fyawgybj.eastus2-01.azurewebsites.net/

### Part 1: Study Information

**Study Goal:**

The primary goal of this human-computer interaction study is to evaluate the effectiveness and user experience of a new language model (LLM)-aided tool for labeling irAEs, compared to the current standard process.

**Study Design:**

- **Two Sessions:** You will participate in two separate labeling sessions.
- **Task:** In each session, you will be asked to label a maximum of 10 clinical notes within a one-hour time limit.
- **Conditions:**
    - **Control Session:** You will use a standard interface, reading the clinical notes directly and inputting the required information into a Microsoft Excel document.
    - **LLM-Aided Session:** You will use a specialized interface that displays the clinical note alongside predictions and supporting evidence generated by a language model.
- **Crossover Design:** Half of the participants will start with the Control session, and the other half will start with the LLM-Aided session. You will switch conditions for your second session.

**Session Recording and Think-Aloud Protocol:**

- During both labeling sessions, your screen activity will be recorded.
- **Purpose:** This helps us manually record your labels if technical difficulties occur.

### Part 2: Annotation Guidelines

#### 1. Introduction and Purpose Statement (Overall Project)

Our ability to understand cancer treatment-related adverse effects, including immune-related adverse events (irAEs), is limited by a lack of data. Cancer treatment side effects are not collected in most cancer registries, and clinical trials are inherently limited in their ability to collect adverse event data. The goal of this project is to develop natural language processing (NLP) methods that can automatically extract information about irAEs in adult cancer patients. The ultimate goal is to use these methods to create real world evidence on irAEs, and to enable technologies to automatically monitor patients' electronic health records (EHRs) for these events.

These guidelines describe specific irAEs we wish to capture in the clinical documents of patients undergoing or completed immunotherapy, and the methodology for annotation. In addition to identifying mentions of disorders that may be irAEs in documents, each irAE will have certain attributes to further enrich our datasets.

Human annotations were used to develop our LLM tool that can "read" clinical documents, identify toxicities of interest, and extract them from the texts. We are now validating if this model can help humans to do this faster and more accurately than on their own.

Annotators should carefully read these guidelines in their entirety prior to starting annotations. Please adhere as closely to these guidelines, even in cases that you disagree with definitions or rules provided. This is important to ensure annotation quality and consistency.

#### 2. Annotation Task Overview

The annotation task is to provide each document with labels that provide information about the presence of irAEs, along with descriptive attributes. **Do not use information from other notes on the same patient to arrive at a label.**

Please note that the irAEs and their attributes may not be present in the note as described verbatim below. There are varied natural language expressions of each class, so please use best judgement in determining if the class or attribute is present.

**Priority Disorders:**

We will start labeling only the 6 priority disorders for now:

- Myocarditis (Myo)
- Dermatitis (Derm)
- Thyroiditis (Thyr)
- Hepatitis (Hep)
- Colitis (Col)
- Pneumonitis (Pneum)

It is important to note that these irAEs are umbrella terms for multiple CTCAE terms. For example, an immune-related cough should be labeled as pneumonitis. The term mappings are in **Appendix I: CTCAE term mappings**.

**Context:**

All notes were selected such that the note was written within 12 months of an immunotherapy infusion. You may use this information when determining the irAE labels.

**Important Guiding Principles:**

1. **Label the Differential Diagnosis:** The goal is to label medical events for which the irAE is in the differential diagnosis, not every single isolated sign or symptom that may overlap with an irAE. These irAEs are generally defined by clusters of signs/symptoms that, taken together, are suggestive of the disorder. Often, there will be other diagnoses in the differential at the time the note is written. As long as the irAE is in the differential, you should still label the irAE if it is an unlikely or possible diagnosis, unless there is a clear alternative diagnosis (see point 2 below). Lists of signs and symptoms often seen with each of the 6 priority irAEs, as well as common diagnostic procedures, are provided below (Section 5a: irAE Type) – these can guide whether an irAE is in the differential, even if the exact term is not used in the note.
2. **Exclude Clear Alternative Diagnoses:** If the presentation is similar to an irAE but there is clearly another diagnosis that explains the presentation, and it is stated as such in the note, do not label the irAE as present.
    - *Example:* If a patient is already diagnosed with an infectious pneumonia and the note says that the pneumonia is the cause of shortness of breath or fever, no need to label dyspnea and fever or any associated context spans – the dyspnea and fever are not describing a pneumonitis event.
    - *Example:* If a patient has chronic shortness of breath that is clearly attributed to longstanding COPD, no need to label it – the shortness of breath is not describing a pneumonitis event.
    - *Example:* If a patient is having chest pain but it is clearly because of coronary artery disease, no need to label it – the chest pain is not describing a myocarditis event.

**3. Label Definitions**

The table below describes the labels and their value sets for this project. For each irAE type (indicated by the abbreviation suffix like _Pneum, _Col, etc.) and temporal aspect (Current vs. Past), we label its presence/absence, CTCAE grade, attribution, and certainty. The **Attributes** section (Section 5 below) provides details for the irAE type, Grade, Attribution, and Certainty labels. Please read through these details carefully and reference them as you annotate.

| **Label** | **Name** | **Description** | **ValueSet** |
|---|---|---|---|
| CurrentType_* | Current irAE Type | At the time the note is written, is the patient currently experiencing the irAE? Label yes if the irAE is in the differential, even if it is not certain. Details: irAE Type | 1=yes; blank = no |

| Label | Name | Description | ValueSet |
|---|---|---|---|
| CurrentGrade_* | Current Grade | If yes to CurrentType, what is the current CTCAE grade of that irAE? Details: Severity | 1=grade 1; 2=grade 2; 3=grade 3; 4=grade 4; 5=grade 5 |
| CurrentAttr_* | Current Attribution | If yes to CurrentType, what is the Attribution? Details: Attribution | 1 = IO; blank = not IO |
| CurrentCert_* | Current Certainty | If yes to CurrentType & CurrentAttr=1, what is the explicit certainty that the event is due to immunotherapy? Details: Certainty | 1 = unlikely; 2 = Possible; 3 = Likely; 4 = Certain; blank = no explicit certainty described |
| PastType_* | Past irAE Type | Did the patient have that irAE type in the past? Label yes if the irAE was in the differential and no other clear alternative diagnosis was determined, even if it was not certain. Details: irAE Type | 1=yes; blank = no |
| PastMaxGrade_* | Past Maximum Grade | If yes to PastType, what was the maximum grade (not counting the current grade, if relevant)? Details: Severity | 1=grade 1; 2=grade 2; 3=grade 3; 4=grade 4; 5=grade 5 |
| PastAttr_* | Past Attribution | If yes to PastType, what is the Attribution? Details: Attribution | 1 = IO; blank = not IO |
| PastCert_* | Past Certainty | If yes to PastType & PastAttr=1; what is the explicit certainty that the event is due to immunotherapy? Details: Certainty | 1 = unlikely; 2 = Possible; 3 = Likely; 4 = Certain; blank = no explicit certainty described |

**Each label is appended by an abbreviation for the irAE type in question: Pneum, Col, Derm, Hep, Thyr, Myo*

**4. Temporal Aspects (Current vs. Past irAEs)**

There are different ways irAEs may be described over time which largely break down into the following buckets:

- "The patient previously had immunotherapy-related colitis treated with steroids. The patient now presents with recurrence of the colitis." The current event would be labeled with IO = yes and Certainty = certain because it is clearly described as a recurrence of the past event where clearly IO = yes and Certainty = certain:
    - CurrentType_col = 1 (yes)
    - CurrentGrade_col = 1 (max inferrable grade based on this text)
    - CurrentAttr_col = 1 (IO)
    - CurrentCert_col = 4 (Certain)
    - PastType_col = 1 (yes)
    - PastMaxGrade_col = 2 (inferred from steroid treatment)
    - PastAttr_col = 1 (IO)
    - PastCert_col = 4 (Certain)
- "The patient previously had immunotherapy-related colitis treated with steroids. The patient now presents with possible recurrence of colitis." The word recurrence makes it clear that the current colitis described refers to the past colitis. The past colitis is Attribution = yes, and so the current colitis inherits this same attribution. However, the Certainty is different for the past and present event.
    - CurrentType_col = 1 (yes)
    - CurrentGrade_col = 1 (max inferrable grade based on this text)
    - CurrentAttr_col = 1 (IO)
    - CurrentCert_col = 2 (Possible)
    - PastType_col = 1 (yes)
    - PastMaxGrade_col = 2 (inferred from steroid treatment)
    - PastAttr_col = 1 (IO)
    - PastCert_col = 4 (Certain)
- "Previously admitted for pembro-related pneumonitis… now is admitted for dyspnea due to pneumonia vs inflammatory process such as pneumonitis (2/2 ?too quick discontinuation of steroid taper for prior pneumonitis.)"
    - CurrentType_Pneum = 1 (yes)
    - CurrentGrade_Pneum = 3 (because admitted to the hospital for this event)
    - CurrentAttr_Pneum = 1 (IO)
    - CurrentCert_Pneum = 2 (Possible) because the prior pneumonitis is attributed certainly to pembrolizumab, but this event is only possibly a recurrence of the pembro-induced pneumonitis (because of the "vs"). We know that the possible pneumonitis is felt to be a recurrence of the prior pembro-related pneumonitis because of "2/2 ?too quick discontinuation of steroid taper for prior pneumonitis".
    - PastType_Pneum = 1 (yes)
    - PastMaxGrade_Pneum = 3 (inferred from prior admission)
    - PastAttr_Pneum = 1 (IO)
    - PastCert_Pneum = 4 (Certain)

#### 5. Attributes

##### a. irAE Type

irAE type is just as it sounds – it is the type of irAE. Descriptions of the irAEs below are adapted from the A151804 protocol. For the first phase of the project, we are only identifying the following irAEs: myocarditis, pneumonitis, dermatitis, hepatitis, colitis, thyroiditis. We may extend to other irAEs in the future. It is important to note that these irAEs are umbrella terms for multiple CTCAE terms. For example, immune-related cough should be labeled as pneumonitis. The term mappings are in **Appendix I: CTCAE term mappings**.

**Important Grading Rules:**

- **Always assign a grade** if the irAE Type is present (value=1).
- **Default to Grade = 1** if there is otherwise not enough information to give a higher grade. For example, if all we know about the irAE is that it caused the IO to be discontinued or put on hold, Grade = 1.
- **Grade 3:**
    - If someone is admitted to the hospital due to the irAE, then Grade = 3.
    - If someone is admitted to the hospital for something else but also has an irAE, and the irAE is delaying discharge from hospital, Grade = 3.
- **Grade 5:** Death due to the irAE.

**c. Attribution**

The Attribution property indicates whether the irAE is **explicitly attributed to immunotherapy** by the note author. Determine Attribution based only on explicit documentation by the note writer that the irAE is caused by an immunotherapy or not.

- **IO (Value=1):** The Disorder is explicitly described as being caused by immunotherapy by the note author.
    - Mentions of "drug-related" only counts as IO if it is clear that the "drug" refers to the IO – note that many drugs can cause inflammatory adverse events.
    - If it is explicitly stated that an IO was discontinued due to the event, then that counts as attribution = IO because this means the clinician linked this event to the IO.
    - *Examples:*
        - "The patient had immune-related nephritis"
        - "Patient recently started on nivolumab… presenting with drug-related colitis due to his recent IO treatment".

        - “Pembro was discontinued due to pneumonitis”
- **Not IO (Value=blank/no value):** The disorder is not explicitly attributed to IO. **This is the default label.**
    - *Example:*
        - “Patient had a rash after 2nd infusion of nivolumab” - This is Not IO even though the timing seems like it should be due to immunotherapy, because the clinician does not explicitly say that the rash is caused by immunotherapy.

**d. Certainty**

Certainty refers to **explicit mentions of the clinician’s certainty** that the event is related to immunotherapy. It does not refer to the annotator’s certainty. Terms here mirror the terminology used in the Naranjo scale (https://www.ncbi.nlm.nih.gov/books/NBK548069/). Only add a label if there is an explicit certainty level described; otherwise, leave it blank. This label is only applicable if Attribution is IO (Value=1).

- **None (No explicit certainty described) (Value=blank/no value):** The event is explicitly attributed to immunotherapy (Attribution=IO), but there is no described level of certainty that the event is related to immunotherapy. **This is the default label for Certainty.**
    - *Example:*
        - “Pembro was discontinued due to pneumonitis” (Attribution=IO, Certainty=None/blank) - While this suggests the provider thought the event was due to immunotherapy, the certainty is not clear as they may have discontinued even though there was uncertainty.
    - *Example:*
        - “I will prescribe steroids for pneumonitis.” (Attribution=Not IO, Certainty=None/blank) - No explicit attribution or certainty.

**6. Notes and Edge Cases**

a. **Certainty Complexity:** Statements of certainty (or lack thereof) may complicate labeling and may require the annotator’s best judgement. Please use your best judgement and refer to the Certainty definitions above.
b. **Contradictory Information:** It is possible a document may contradict itself. In these cases, please use your best judgement but in general preference the positive/present/more severe labels.
c. **Negated Entities:** Do not label descriptions of a disorder *not* being present. For example, “The patient has no pneumonitis.” should not be labeled.
d. **Copy-pasted Radiology Reports:** Radiology reports or sections of reports are often copy-pasted into other clinical notes. In some cases, these describe an irAE. If the label(s) based on the radiology report disagrees with the clinical note label(s), preference the clinical note label(s). For CTCAE Grade 1 events, there may be only mention of the irAE being present or possibly present in the radiology report. In these cases, you should label according to the radiology report even if the irAE is not mentioned elsewhere in the note.
e. **Laboratory Values:** Do not use lab values in isolation to label irAEs. They should be considered in the context of other signs, symptoms, and clinical assessments mentioned in the note.

### Part 3: Labeling Interface Overview

You will use two different interfaces during the study:

1. **Control Interface:** This involves reading the clinical notes provided (likely as text files or directly on a website) and manually entering your labels into a structured Microsoft Excel document.
2. **LLM-Aided Interface:** This is a specialized web-based tool that will display the clinical note alongside potential irAE mentions, attributes, and supporting text evidence suggested by a language model. You will review these suggestions, correct them as needed, and finalize the labels within this interface.

*Note: More specific instructions on navigating and using each interface will be provided during the training session or as separate guides.*

#### *3.1 Control Interface*

In the Control condition, you will:

1. **View the note only.**
   – You will see each clinical note displayed in plain text on a web page, **without** any highlighting or model suggestions.
2. **Record your labels in Google Sheets.**
   – Open the provided Google Sheet (see example_control_csv).
   – For each note, identify and record:
   - **Condition name** (e.g., “colitis,” “hepatitis,” etc.)
   - **Presence** (Yes/No)
   - **Grade** (0–5; enter 0 if absent)
   - **Supporting text** (copy the sentence or phrase that justifies your label)
3. **Proceed note by note.**
   a. Read the full note.
   b. Complete all columns in the Google Sheet for that note.
   c. Click **Next** on the web page to load the next note.
   d. A small pop-up will confirm the note ID and your entries; verify accuracy, then continue.

**Tip:** If you believe a note contains no occurrences of any condition, simply record “No” for presence and “0” for grade. Leave the supporting-text field blank.

#### *3.2 LLM-Aided Interface*

In the LLM-Aided layout (here), you will:

1. **See the split-screen layout.**
   a. **Left pane:** The full clinical note, with any model-identified “evidence spans” highlighted in yellow.
   b. **Right pane:** A list of the 6 target conditions, each showing:
      i. A filled dot if the model detected evidence (empty dot if none)
      ii. Model-suggested **Grade (current/past)**
      iii. A **“View Evidence”** link that attempts to scroll the left pane to the highlighted passage
2. **Review and adjust model suggestions.**
   **a. To accept a suggestion:**

i. Click **“Use label”** next to the Grade or Presence field—this populates the form below.

**b. To add a new label or correct the suggestion:**

i. Click **“Add label”**, then manually select the correct condition, grade, and supporting text.

**3. Complete the label form.**

a. The form at the bottom lets you:

i. Confirm the **Condition**

ii. Select **Presence** (Yes/No)

iii. Enter **Grade** (0–5; default 0 if none)

iv. Add the Attribution and Certainty

v. If you want to add a comment, this is not required and is not a substitute for submitting a label.

b. Only the active tab’s condition will be added when you click **add Label**.

c. If **all grades are 0** (no finding), you do **not** need to submit—absence defaults to grade 0.

**4. Navigate through notes.**

a. After submitting, click **Next** to proceed.

b. A summary dialog will display all six conditions and your finalized labels—review, then confirm to save.

**Note:** “View Evidence” relies on an exact text match. If the highlight link fails, scroll manually or use the search bar to locate the suggested span.

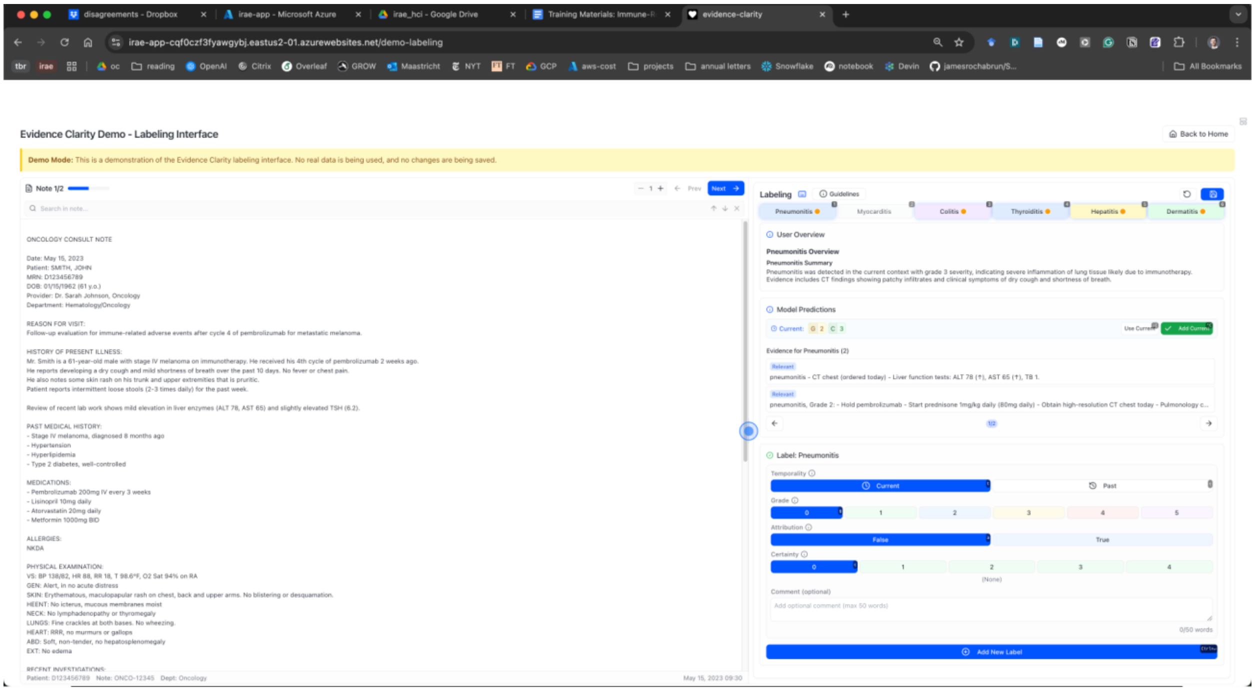

#### Conclusion and Next Steps

Thank you for carefully reviewing these training materials. Please come prepared with any questions you may have for the upcoming training session. Your participation is crucial for helping us understand how best to leverage technology for identifying and understanding treatment-related adverse events.

### Appendix I: CTCAE term mappings

Alliance A151804 irAEs of interest, and related AEs listed in CTCAE v5, include:

| irAE | CTCAE MedDRA Code | CTCAE v5 terms |
|---|---|---|
| Myocarditis | 10003586<br>10003658<br>10003662<br>10003673<br>10007515<br>10007541<br>10010276<br>10019279<br>10069501<br>10027786<br>10027787<br>10028596<br>10028606<br>10034040<br>10034474<br>10053565<br>10034484<br>10058597<br>10040639<br>10042604<br>10047281<br>10047290<br>10047302<br>10007612<br>10007613<br>10014383 | Asystole<br>Atrial fibrillation<br>Atrial flutter<br>Atrioventricular block complete<br>Cardiac arrest<br>Cardiac disorders – Other, specify<br>Conduction disorder<br>Heart failure<br>Left ventricular systolic dysfunction<br>Mobitz (type) II atrioventricular block<br>Mobitz type I<br>Myocardial infarction<br>Myocarditis<br>Paroxysmal atrial tachycardia<br>Pericardial effusion<br>Pericardial tamponade<br>Pericarditis<br>Right ventricular dysfunction<br>Sick sinus syndrome<br>Supraventricular tachycardia<br>Ventricular arrhythmia<br>Ventricular fibrillation<br>Ventricular tachycardia<br>Cardiac troponin I increased<br>Cardiac troponin T increased<br>Electrocardiogram QT corrected interval prolonged |
| Colitis | 10000081<br>10065747<br>10009887<br>10009998<br>10012727<br>10014893<br>10021328<br>10038064 | Abdominal pain<br>Cecal hemorrhage<br>Colitis<br>Colonic hemorrhage<br>Diarrhea<br>Enterocolitis<br>Ileus<br>Rectal hemorrhage |
| Hepatitis | 10001551<br>10001675<br>10003481<br>10005364 | Alanine aminotransferase increased<br>Alkaline phosphatase increased<br>Aspartate aminotransferase increased<br>Blood bilirubin increased |
| Pneumonitis | 10011224<br>10013963<br>10021143<br>10035742 | Cough<br>Dyspnea<br>Hypoxia<br>Pneumonitis |

| | | |
|---|---|---|
| Dermatitis | 10006556<br>10015218<br>10015277<br>10037868<br>10042033<br>10044223 | Bullous dermatitis<br>Erythema multiforme<br>Erythroderma<br>Rash maculo-papular<br>Stevens-Johnson syndrome<br>Toxic epidermal necrolysis |
| Endocrinopathies | 10001367<br>10020639<br>10020850<br>10062767<br>10021114<br>10043770 | Adrenal insufficiency<br>Hyperglycemia<br>Hyperthyroidism<br>Hypophysitis<br>Hypothyroidism<br>Thyroid stimulating hormone increased |
