## Appendix 5b for "An agentic AI system enhances clinical detection of immunotherapy toxicities: a multi-phase validation study"

**Pre-Study Survey: Clinical Note Labeling for Immunotherapy Adverse Events**

*Thank you for participating in this study. This survey gathers information about your background, experience, and views on AI, particularly in the context of clinical documentation analysis. Your responses will help us understand the context of the study results.*

---

**Section 1: Demographics & Role**

1. **Gender**
2. **Demographics**
3. **What is your name?**
    - [Free Text]
4. **What is your primary clinical role for this study?**
    - Clinical Research Coordinator (CRC)
    - Nurse
    - Other (Please specify): [Free Text]

---

**Section 2: Role-Specific Experience**

**(IF CRC selected)**

5. **How many years have you been a CRC?**
    - [Number] years
6. **How many years have you worked in oncology research?**
    - [Number] years
7. **How many years have you been involved in studies using immunotherapy agents?**
    - [Number] years
8. **How would you rate your experience with identifying and documenting Adverse Events (AEs), specifically Immunotherapy-Related Adverse Events (irAEs), from clinical notes or patient records?**
    - None
    - Slight (Basic familiarity)
    - Moderate (Regularly perform this task)
    - Extensive (Significant part of my role, high volume)
    - Expert (Train others, deep expertise)
9. **How familiar are you with CTCAE (Common Terminology Criteria for Adverse Events) grading?**
    - Not familiar
    - Slightly familiar (Have heard of it / used it rarely)
    - Moderately familiar (Use it sometimes / understand the basics)
    - Very familiar (Use it regularly / comfortable with most grades)

- Extremely familiar (Use it constantly / can grade complex events confidently)

**(IF NURSE selected)**

10. **How many years have you been practicing as a Nurse?**
    - [Number] years
11. **How many years have you worked in an oncology setting?**
    - [Number] years
12. **How many years have you been involved in the care of patients receiving immunotherapy?**
    - [Number] years
13. **How would you rate your experience with identifying, documenting, and/or managing Adverse Events (AEs), specifically Immunotherapy-Related Adverse Events (irAEs), based on clinical notes or patient interactions?**
    - None
    - Slight (Basic familiarity)
    - Moderate (Regularly involved in this process)
    - Extensive (Significant part of my role, manage complex cases)
    - Expert (Lead protocols, educate others, deep expertise)
14. **How familiar are you with CTCAE (Common Terminology Criteria for Adverse Events) grading?**
    - Not familiar
    - Slightly familiar (Have heard of it / used it rarely)
    - Moderately familiar (Use it sometimes / understand the basics)
    - Very familiar (Use it regularly / comfortable with most grades)
    - Extremely familiar (Use it constantly / can grade complex events confidently)

---

**Section 3: Experience and Views on AI & Automation (Everyone)**

15. **Do you have prior experience with Artificial Intelligence (AI) in general (outside of medicine, e.g., chatbots, recommendation systems)?**
    - None
    - Slight
    - Moderate
    - Very
    - Extreme
16. **Do you have prior experience with any AI tools used *within* a clinical or healthcare setting?**
    - None
    - Slight (e.g., basic EHR alerts)
    - Moderate (e.g., used AI diagnostic aids, automated charting tools)
    - Very (e.g., regularly use multiple AI tools in practice)
    - Extreme (e.g., involved in developing/implementing clinical AI)
17. **Do you conduct research involving AI?**

- None
- Slight (e.g., user in an AI study)
- Moderate (e.g., contribute data or clinical insights to AI projects)
- Very (e.g., actively involved in designing/running AI research)
- Extreme (e.g., principal investigator on AI grants/projects)

18. **How much trust do you generally have in AI systems for performing non-medical tasks?**

- None
- Slight
- Moderate
- Very
- Extreme

19. **How much trust do you specifically have in Large Language Models (LLMs, like ChatGPT, Bard, etc.) for tasks involving clinical information?**

- None
- Slight
- Moderate
- Very
- Extreme

20. **How often do you currently use any tools for automated *information extraction* or *summarization* from clinical notes in your practice (e.g., NLP features in EHR, specific research tools)?**

- Never
- Rarely
- Sometimes
- Often
- Very Often / Routinely

21. **How often do you use Large Language Models (LLMs like ChatGPT, Bard, clinical specific variants, etc.) for any *work-related* tasks (e.g., drafting emails, searching information, summarizing text)?**

- Never
- Rarely
- Sometimes
- Often
- Very Often / Routinely

22. **How often do you use any AI-based automation or productivity tools (including but not limited to LLMs or information extraction) in your clinical practice or research work?**

- Never
- Rarely
- Sometimes
- Often
- Very Often / Routinely

---

**Section 4: Perspectives on AI/LLMs in Clinical Note Analysis (Everyone)**

23. **Do you believe AI/LLM tools have the potential to *benefit* clinical practice and medicine generally in the long term?**

- Strongly Disagree
- Disagree
- Neutral
- Agree
- Strongly Agree

24. **Do you believe AI/LLM tools have the potential to *harm* clinical practice and medicine generally in the long term?**

- Strongly Disagree
- Disagree
- Neutral
- Agree
- Strongly Agree

25. **What impact do you think AI/LLM tools will have on accuracy and quality compared to manual review?**

- Significant harm
- Moderate harm
- Neutral
- Moderate benefit
- Significant benefit

26. **What impact do you think AI/LLM tools will have on efficiency (speeding up the process) of identifying, grading, and attributing irAEs from clinical notes, compared to manual review?**

- Significant harm
- Moderate harm
- Neutral
- Moderate benefit
- Significant benefit

27. **How confident are you that current Large Language Models (LLMs) can reliably and accurately perform complex clinical interpretation tasks like irAE identification, grading, and attribution based *solely* on unstructured clinical notes?**

- Not at all confident
- Slightly confident
- Moderately confident
- Very confident
- Extremely confident

**Section 5: Additional Comments**

28. **Do you have any additional comments about your experience or expectations regarding AI/LLMs in the context of clinical documentation analysis or irAE identification?**
    - [Free Text]

---

**End of Survey - Thank You!**

---
