## Appendix 5c for "An agentic AI system enhances clinical detection of immunotherapy toxicities: a multi-phase validation study"

**Post-Study Survey: Clinical Note Labeling for Immunotherapy Adverse Events**

*Thank you for completing the study tasks. This survey gathers your feedback on the experience with both the manual labeling process and the AI/LLM assistance tool. Your responses are crucial for evaluating the tool and process.*

---

**Section 1: Experience with the Control (Manual) Interface**

*This section asks about your experience using the interface where you performed the labeling task **WITHOUT** AI/LLM assistance.*

1. **Please rate your confidence in the accuracy of the labels you assigned using the *Control (Manual)* interface:**
    - Statement: I felt confident in the accuracy of the labels I assigned using this interface.
        1. Strongly Disagree
        2. Disagree
        3. Neutral
        4. Agree
        5. Strongly Agree
2. **System Usability Scale (SUS) - Control (Manual) Interface:** *Please rate the following statements for the Control (Manual) interface based on your experience.* (Scale: 1-Strongly Disagree, 2-Disagree, 3-Neutral, 4-Agree, 5-Strongly Agree)
    - i. I think that I would like to use this system frequently. [1 2 3 4 5]
    - ii. I found the system unnecessarily complex. [1 2 3 4 5]
    - iii. I thought the system was easy to use. [1 2 3 4 5]
    - iv. I think that I would need the support of a technical person to be able to use this system. [1 2 3 4 5]
    - v. I found the various functions in this system were well integrated. [1 2 3 4 5]
    - vi. I thought there was too much inconsistency in this system. [1 2 3 4 5]
    - vii. I would imagine that most people would learn to use this system very quickly. [1 2 3 4 5]
    - viii. I found the system very cumbersome to use. [1 2 3 4 5]
    - ix. I felt very confident using the system. [1 2 3 4 5]
    - x. I needed to learn a lot of things before I could get going with this system. [1 2 3 4 5]
3. **Please rate your perceived efficiency using the *Control (Manual)* interface:**
    - Statement: I felt efficient while using this interface to complete the labeling task.
    - Scale: Strongly Disagree - Disagree - Neutral - Agree - Strongly Agree
4. **Fatigue- Control (Manual) Interface:**
    - Statement: This was a manageable amount of notes for 1 hour.
    - Scale: Strongly Disagree - Disagree - Neutral - Agree - Strongly Agree
5. **Overall Satisfaction - Control (Manual) Interface:**
    - Statement: Overall, I was satisfied with my experience using this interface for the task.
    - Scale: Strongly Disagree - Disagree - Neutral - Agree - Strongly Agree
6. **Additional Comments - Control (Manual) Interface:**

- Do you have any additional comments or feedback about the *manual* labeling process you completed using this interface?
- [Free Text]

---

**Section 2: Experience with the AI Assisted Interface**

*This section asks about your experience using the interface where you performed the labeling task* ***WITH*** *AI/LLM assistance/suggestions.*

7. **Please rate the helpfulness of the AI suggestions:**
    - Statement: The AI suggestions provided by the interface were helpful in identifying and labeling the relevant information.
    - Scale: Strongly Disagree - Disagree - Neutral - Agree - Strongly Agree
8. **Please rate your confidence in the accuracy of the labels you assigned using the *AI-Assisted* interface (after your review and edits):**
    - Statement: I felt confident in the accuracy of the labels I assigned using this interface.
    - Scale: Strongly Disagree - Disagree - Neutral - Agree - Strongly Agree
9. **System Usability Scale (SUS) - AI-Assisted Interface:** *Please rate the following statements for the AI/LLM-Assisted interface based on your experience.* (Scale: 1-Strongly Disagree, 2-Disagree, 3-Neutral, 4-Agree, 5-Strongly Agree)
    - i. I think that I would like to use this system frequently. [1 2 3 4 5]
    - ii. I found the system unnecessarily complex. [1 2 3 4 5]
    - iii. I thought the system was easy to use. [1 2 3 4 5]
    - iv. I think that I would need the support of a technical person to be able to use this system. [1 2 3 4 5]
    - v. I found the various functions in this system were well integrated. [1 2 3 4 5]
    - vi. I thought there was too much inconsistency in this system. [1 2 3 4 5]
    - vii. I would imagine that most people would learn to use this system very quickly. [1 2 3 4 5]
    - viii. I found the system very cumbersome to use. [1 2 3 4 5]
    - ix. I felt very confident using the system. [1 2 3 4 5]
    - x. I needed to learn a lot of things before I could get going with this system. [1 2 3 4 5]
10. **Please rate your perceived efficiency using the *AI-Assisted* interface:**
    - Statement: I felt efficient while using this interface to complete the labeling task.
    - Scale: Strongly Disagree - Disagree - Neutral - Agree - Strongly Agree
11. **Overall Satisfaction - AI-Assisted Interface:**
    - Statement: Overall, I was satisfied with my experience using this interface for the task.
    - Scale: Strongly Disagree - Disagree - Neutral - Agree - Strongly Agree
12. **Fatigue Control - AI-Assisted Interface:**
    - Statement: This was a manageable amount of notes for 1 hour.
    - Scale: Strongly Disagree - Disagree - Neutral - Agree - Strongly Agree
13. **Additional Comments - AI-Assisted Interface:**

- Do you have any additional comments or feedback about the *LLM-assisted* labeling process you completed using this interface?
- [Free Text]

---

**Section 3: Comparison and Impact**

*Now, please compare your experiences between the two approaches.*

14. **Compared to performing the task manually, how did using the AI assistance tool affect the *perceived overall quality* (accuracy, completeness) of your final irAE labels?**
    - Significantly Hurt Quality
    - Slightly Hurt Quality
    - No Difference
    - Slightly Improved Quality
    - Significantly Improved Quality
15. **Compared to performing the task manually, how did using the AI assistance tool affect the *efficiency (speed)* of your labeling process?**
    - Significantly Slowed Me Down
    - Slightly Slowed Me Down
    - No Difference
    - Slightly Sped Me Up
    - Significantly Sped Me Up
16. **How mentally demanding did you find the labeling task *with* AI assistance compared to *without* it?**
    - Much More Demanding with AI
    - Slightly More Demanding with AI
    - About the Same Demanding
    - Slightly Less Demanding with AI
    - Much Less Demanding with AI
17. **Overall, which condition did you *prefer* for completing the irAE labeling task?**
    - Strongly Prefer Manual Labeling
    - Prefer Manual Labeling
    - Neutral / No Preference
    - Prefer AI-Assisted Labeling
    - Strongly Prefer AI-Assisted Labeling

---

**Section 4: Changes in Perception, Trust, and LLM Capability**

18. **After participating in this study, how *confident* are you now that** AI **can reliably assist with complex clinical tasks like adverse event identification, grading, and attribution from notes?**
    - Not at all confident
    - Slightly confident
    - Moderately confident

- Very confident
- Extremely confident

19. **Did this experience change your overall perception of the potential *value* (benefit vs. risk) of using AI/LLMs for analyzing clinical notes for adverse events?**
    - Yes my perception became significantly more negative.
    - Yes my perception became slightly more negative.
    - No my perception did not change significantly.
    - Yes my perception became slightly more positive.
    - Yes my perception became significantly more positive.
20. **Do you think this experience will change anything about how you approach *manual* irAE identification, grading, or attribution in your regular work?**
    - Definitely Not
    - Probably Not
    - Unsure / Maybe Slightly
    - Probably Yes
    - Definitely Yes
    - *(Optional Follow-up):* If yes, please briefly describe how: [Free Text]
21. **Based on your experience with the** AI **tool in this study, how would you rate its potential to perform the irAE labeling task compared to human roles?**
    - Significantly worse than a minimally trained person; not useful.
    - Requires extensive correction; potentially useful for basic identification but not grading/attribution.
    - Comparable to a trained CRC for identification/basic grading, but needs expert review.
    - Comparable to an experienced Nurse/CRC/Physician for most aspects, requiring moderate oversight.
    - Potentially better/more consistent than human reviewers for specific aspects, requiring minimal oversight.
22. **Based on your experience, how much human oversight or review do you think would be necessary if *this specific*** AI ***tool (as experienced)*** **were used today in a real-world setting for irAE documentation?**
    - Tool is not reliable enough for use, requires complete manual re-work.
    - Extensive review required for all cases and all labels.
    - Moderate review needed for most cases, focusing on complex events or higher grades.
    - Some spot-checking required, mainly for quality assurance.
    - Minimal oversight needed (Tool is highly reliable).

---

**Section 5: General Views on AI (Post-Experience)**

*We are asking these general questions again to see if the experience influenced your broader views.*

23. **Do you believe AI/LLM tools have the potential to *benefit* clinical practice and medicine generally in the long term?**
    - Strongly Disagree
    - Disagree

- Neutral
- Agree
- Strongly Agree

24. **Do you believe AI/LLM tools have the potential to *harm* clinical practice and medicine generally in the long term?**

- Strongly Disagree
- Disagree
- Neutral
- Agree
- Strongly Agree

---

**Section 6: Overall Feedback and Suggestions**

25. **What aspect(s) of the** AI **assistance tool did you find *most useful* or helpful during the task?**

- [Free Text]

26. **What aspect(s) of the** AI **assistance tool were *most frustrating, least useful, or inaccurate*?**

- [Free Text]

27. **Do you have any specific suggestions for improving the** AI **tool, its interface, or the way it presents information for this task?**

- [Free Text]

28. **Do you have any other comments about the study tasks, the instructions, or your overall experience?**

- [Free Text]

---

**End of Survey - Thank You for Your Valuable Feedback!**
